# Low Responsiveness of Machine Learning Models to Critical or Deteriorating Health Conditions

**DOI:** 10.1101/2024.09.25.24314400

**Authors:** Tanmoy Sarkar Pias, Sharmin Afrose, Moon Das Tuli, Ipsita Hamid Trisha, Xinwei Deng, Charles B. Nemeroff, Danfeng (Daphne) Yao

## Abstract

**Background:** Machine learning (ML) based mortality prediction models can be immensely useful in intensive care units. Such a model should generate warnings to alert physicians when a patient’s condition rapidly deteriorates, or their vitals are in highly abnormal ranges. Before clinical deployment, it is important to comprehensively assess models’ ability to recognize critical patient conditions.

**Methods:** We develop testing approaches to systematically assess machine learning models’ ability to respond to serious medical emergencies by generating such conditions. We evaluated multiple machine learning models trained on four different datasets from two different clinical prediction tasks and evaluated ten different machine learning models including three resampling techniques.

**Results:** We identified serious deficiencies in the models’ responsiveness, i.e., the inability to recognize severely impaired medical conditions or rapidly deteriorating health. For in-hospital mortality prediction, the models tested using our synthesized cases fail to recognize 66% of the test cases involving injuries. In some instances, the models fail to generate adequate mortality risk scores for all test cases. Our testing methods identified similar kinds of deficiencies in the responsiveness of 5-year breast and lung cancer prediction models.

**Conclusion:** Using generated test cases, we found that statistical machine-learning models trained solely from patient data are grossly insufficient and have many dangerous blind spots. Despite their overall performance metrics, most ML models we tested failed to respond adequately to critically ill patients. Our proposed testing framework acts as a critical bridge between machine learning model development and clinical deployment, ensuring reliability and safety through rigorous evaluation.

**Plain Language Summary:** Machine learning models are increasingly used in healthcare to predict patients’ death risk or cancer survivability. These models could help doctors identify patients with worsening condition and take action to save lives. We developed a new method to test how well these models respond to severe health emergencies by creating life-threatening scenarios. We found most models failed to respond adequately to critical health events. For example, models missed 66% of cases involving serious injuries. This research emphasizes that current machine learning models have major limitations and could be dangerous if used in hospitals without thorough testing. Our testing framework can help improve these models to ensure they are safe and reliable before being used in real-life medical settings.

## Introduction

Artificial intelligence (AI) machine learning technologies are rapidly made available for incorporation into clinical workflows. The Food Drug Administration (FDA) authorized the first autonomous AI diagnostic system in 2018, which is for detecting diabetic retinopathy^1^. Reports also showed that hospitals have adopted machine learning models for clinical uses^2^, e.g., Sepsis Watch at Duke University Hospital reduced inpatient mortality from 9.6% to 8.1%^3^. AI models are also deployed to predict surgery time in hospitals^4^.

For medical applications, mispredictions of machine learning (ML) models may have serious consequences. For example, missed detection (i.e., false negatives) in-hospital mortality prediction or 5-year cancer prognosis^5^ may cause death or underestimate patients’ risks. A widely deployed early warning system for sepsis in U.S. hospitals, the Epic Sepsis Model, was found to give poor prediction performance (AUC-ROC 0.63) in a study involving 27,697 patients^6^. Many factors contribute to mispredictions. It is widely recognized that biased training data cause prediction errors, especially for minority patients in the minority disease class, e.g., Black patients who die in the hospital^5,7^. The sole use of whole population-wide metrics may also mask the deficiencies in the prediction accuracy of minority-class patients, further exacerbating the problem^5^. Noisy training images may also interfere with the model’s learning process, as shown in a skin cancer application^8^. Similar accuracy concerns also exist in other critical AI applications, such as self-driving vehicles, which have resulted in fatalities^9^ and severe injuries^10^.

For mortality prediction, it is important to measure whether or not ML models can promptly respond to deteriorating patients’ conditions. Systematically testing medical AI machine learning models before clinical adoption can help reveal the prediction deficiencies, motivate model corrections, and improve their trustworthiness^11^. Exhaustive testing is both unnecessary and impossible in most medical AI applications, due to the immense complexity of the problem space. Thus, it is crucial to develop strategic testing approaches focusing on the most critical conditions.

The current ML testing practice is very limited in terms of *i)* the coverage of minority class cases, *ii)* responsiveness to disease conditions, and *iii)* testing time-series cases. **First**, existing testing is largely restricted to a small percentage (10-15%) of the existing dataset, i.e., test set, as the majority of the data is reserved for training. Because data imbalance in medicine is common, a typical test set likely has a low coverage of various critical medical conditions and minority prediction class cases. For in-hospital mortality prediction, the minority prediction class is the death cases, e.g., 13.5% are death cases and the majority (86.5%) of the patients do not die after staying in hospital. Even with cross-validation and bootstrapping, the test set is largely limited to the original data. During clinical deployment, new patient conditions may occur out of the distribution of the test set. **Second**, a medical AI model’s responsiveness to specific disease conditions needs to be evaluated. A typical machine learning model, prioritizing majority class samples, may underestimate mortality risks and fail to produce high enough mortality risk scores for critically ill patients^5^. For example, we found that a model trained on the MIMIC-III benchmark^12^ gives a recall of 88.7% for non-death cases (majority prediction class), but 57% recall for death cases (minority prediction class). Thus, testing needs to consider this disparity and prioritize underrepresented disease conditions for evaluation. **Third**, time series data is pervasive in medicine.

However, methods for systematically assessing ML models for time series are missing. Generative technologies have been proposed to produce curated manmade images for self-driving vehicles^13,14^. However, image-based solutions do not address the unique temporal challenge in time series applications.

In this work, we develop systematic approaches for generating new test cases beyond the original dataset to assess the responsiveness of machine learning models to critical health conditions that may occur in clinical settings. Our test case generation is guided by domain knowledge and medical experts. Our experiments involve binary classification tasks, including time-series-based in-hospital mortality prediction and 5-year breast and lung cancer survivability prognosis (Figure 1). We develop multiple methods for generating high-risk test cases that do not exist in the training data or are underrepresented in the training set. We also conduct interviews with medical experts to obtain their estimated risks on some of the generated test cases. Our work reveals alarming prediction deficiencies of machine learning models and points out that ML responsiveness is an important aspect of trustworthiness in digital health.

**Figure 1:**
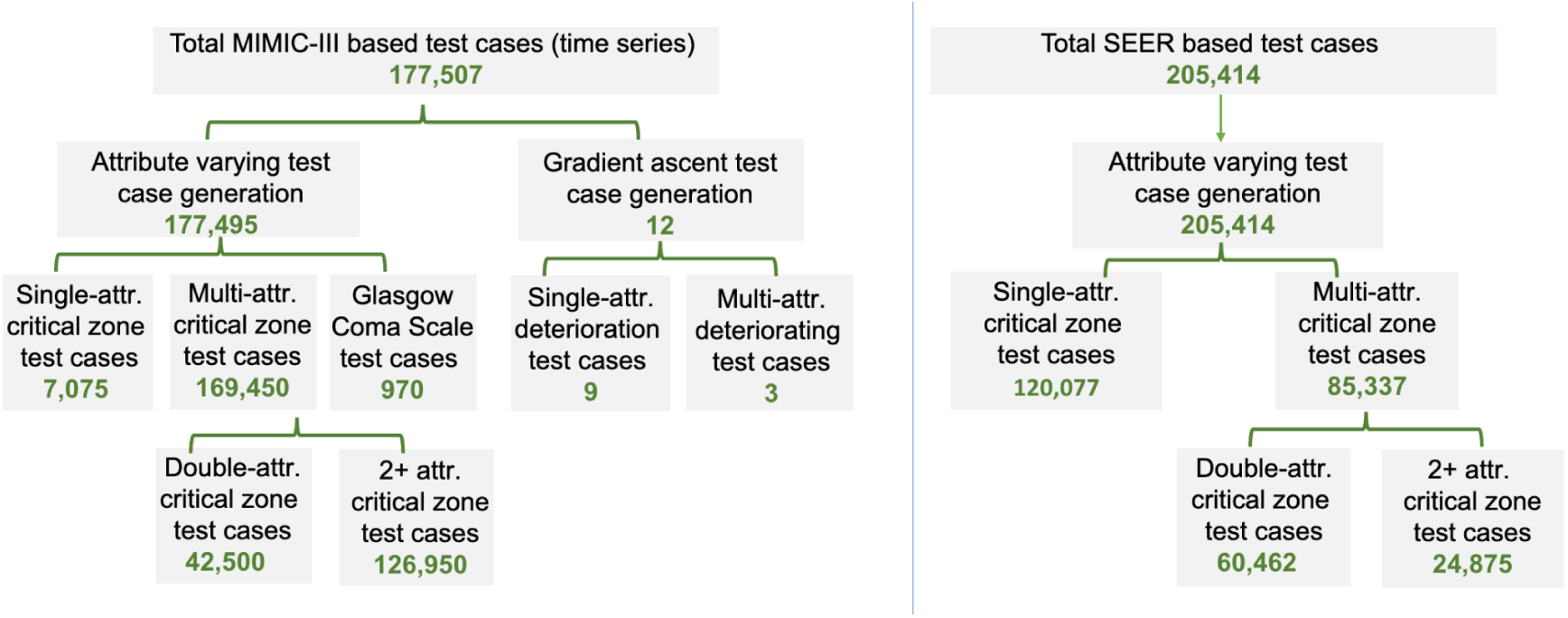
Left side illustrates the generated test case of each category for testing in-hospital mortality risk prediction models trained on MIMIC III or eICU dataset. Right side represents the generated test cases to test 5-year breast cancer survivability (BCS) prediction models. The SEER lung cancer survivability (LCS) models are tested similarly using the single-attribute test cases.

## Methods

### Prediction tasks, datasets, and model selection

Our work aims to test medical machine learning (ML) models for their binary classification accuracy under serious disease conditions. We focus on three binary prediction tasks, namely *i)* 48-hour in-hospital mortality (IHM) risk prediction, *ii)* 5-year breast cancer survivability (BCS) prediction, and *iii)* 5-year lung cancer survivability (LCS) prediction.

The datasets in our study include *i)* a 2019 benchmark^12^ based on the MIMIC III^15,16^ dataset, *ii)* a 2020 benchmark^17^ based on the eICU^18^ dataset, and *iii)* a 2018 reproducibility benchmark^19^ based on the SEER (5-years breast and lung cancer) dataset^19^. The first two datasets contain patients’ 48-hour time series data in critical care units (ICU). Our study excludes clinical free text notes. As with many medical datasets, the MIMIC-III dataset for IHM, containing 21,139 samples, is imbalanced, with 13.2% death cases (Class 1), and 86.8% non-death cases (Class 0). The eICU IHM benchmark dataset contains a total of 30,681 (88.5% for Class 0 and 11.5% for Class 1) samples with similar attributes and time lengths to the MIMIC III benchmark^17^. Supplementary Figure S1 shows the distributions of key attributes of both MIMIC III and eICU datasets. The SEER BCS dataset contains 248,751 patient cases with 56 attributes (7 numerical and continuous features and 49 categorical). In the SEER BCS dataset, 12.7% of cases are death cases (Class 0); the rest are survived cases (Class 1). Supplementary Figure S2 shows the distributions of key attributes. The SEER LCS dataset contains 205,555 cases with 47 features (7 numerical and continuous features and 40 categorical). 84% of patients died in the LCS dataset.

We select ML models that are commonly used in the medical literature for these prediction tasks. Specifically, we select long short term memory (LSTM) as it is widely used for predicting mortality risk in a 48-hour ICU time series dataset— in recent literature^5,20–22^. Similarly, for cancer survivability prediction, we selected multi-layer perceptron (MLP), which was commonly used in analyzing SEER datasets, e.g.,^5,19,23^. In addition, we also evaluated general-purpose ML models commonly seen in medical literature, including XGBoost, AdaBoost, random forest, Gaussian Naive Bayes, and K-nearest Neighbor (KNN). For mortality prediction, we also include CW-LSTM and linear logistic regression models from the benchmark study^12^ and an advanced transformer model.

### Dataset preprocessing

We train machine learning models with benchmark datasets of MIMIC-III^12^, eICU^17^, and SEER breast and lung cancer survivability studies^19^, following the conventional pre-training processing (e.g., encoding, standardization). As MIMIC-III and eICU benchmark datasets contain missing values, we imputed the values that are missing using the most recent observation (within 48 hours) if it exists, otherwise, a value from the normal range of corresponding vitals is mentioned in^12^. Masking was used to indicate whether the vital value was original or imputed. The categorical variables, including binary ones, were encoded using a one-hot vector. The numerical features, such as diastolic blood pressure and glucose level, were converted to their standardized form. After preprocessing, each time-series data point became a 76-by-48 matrix (76 computed features and 48 hours). The processed dataset was used for training and testing neural network-based models such as LSTM and CW-LSTM models. For non-neural network models that cannot directly process time series, we extracted 6 statistical features (mean, min, max, standard deviation, skew, and number of measurements) from various sub-periods (first/last 10%, 25%, 50%, and full 100%). We did not encode the categorical variables, as they contain values with a meaningful scale. The missing values were replaced with mean values computed on the training set and numerical variables were standardized. In total, we obtained 714 features from each 48-hour time series with 17 vitals. The continuous variables were standardized before training. After encoding, the feature vector length of the BCS and LCS datasets became 1,418 and 1,314, respectively.

### Configurations of machine learning models

For in-hospital mortality risk prediction, we utilized the long short term memory (LSTM) model, channel-wise long short term memory (CW-LSTM) model, transformer, logistic regression (LR), AdaBoost, XGBoost, and random forest (RF) models. For 5-year breast cancer survivability (BCS) prediction, we used multi-layer perceptron (MLP), AdaBoost, XGBoost, and random forest models. We utilized the optimal settings of neural network models (i.e., layers, activation, hyperparameters) for each of the tasks from corresponding benchmarks^12,19^. The LSTM model consisted of an input layer (76 dimensions), a masking layer (76 dimensions), a bidirectional LSTM layer (16 dimensions), an LSTM layer (16 dimensions), a dropout layer, and finally a dense layer (1 dimension). In total, the LSTM had 7,569 trainable parameters. The CW-LSTM layer consisted of an input layer (76 dimensions), masking layer (76 dimensions), 17 channel layers (for each 17 input features), 17 bidirectional layers (connected to one of the 17 channels layers), another set of 17 bidirectional layers, a concatenation layer connecting all 17 bidirectional layers, bidirectional layer (64 dimensions), LSTM layer (36 dimensions), dropout layer (64 dimensions), and finally a dense layer (1 dimension). In total, the CW-LSTM model had 153,025 parameters. The size of CW-LSTM’s parameters was 20 times that of LSTM’s. The CW-LSTM model allows independent pre-processing of each variable before combining them. For both LSTM-based models, the optimal hyperparameters are selected using grid search^12^. For example, the batch size, dropout, and time-step are set to 8, 0.3, and 1, respectively. The transformer model consisted of an input layer (76 dimensions), a masking layer (76 dimensions), a positional encoding layer (76 dimensions), 2-3 transformer encoder blocks, a global average pooling layer, a batch normalization layer, a dropout layer (0.3 or 05), a dense layer (32 or 64 units), and finally a dense layer (1 dimension). Each transformer encoder block included a multi-head attention layer with 4 heads (key dimension 76), followed by layer normalization and residual connections. The feed-forward dense layers within each encoder block contained a hidden dimension of 16. In total, the transformer model contains a total of 881,677 parameters (trainable parameters: 293,841, optimizer parameters: 587,684, and non-trainable parameters: 152), larger than both the LSTM and CW-LSTM models. For hyperparameter tuning, grid search was employed to select the best hyperparameters (Supplementary Table 1). The logistic regression (LR) model was from the sklearn library, utilizing the L2 regularization penalty. To prevent overfitting and to enhance the generalization capability of the model, the parameter C is 0.001. This choice of a small C value effectively controls the amount of regularization applied during training. The remaining hyperparameters were left at their default values, following the standard implementation provided by the Python Sklearn library. This model was trained with the standardized training set. The MLP model used for BCS survivability prediction consists of 2 hidden layers, where each hidden layer contains 20 neurons. The hidden layer used Relu as an activation function. Dropout rate of 0.1 after each hidden layer was used to avoid overfitting. The last layer predicted binary labels using the sigmoid activation function. The MLP model contained 28,831 trainable parameters. MLP hyperparameter is empirically selected using grid-search from a list of predefined values such as the number of hidden layers (1, 2, 3, and 4), number of nodes in each layer (20, 50, 100, and 200), and dropout (0, 0.1, 0.2, 0.3, 0.4, and 0.5)^19^. The other models are implemented using Python’s Sklearn library and hyperparameters are tuned using grid search (Supplementary Table 1).

### Model training, threshold tuning, and imbalance correction methods

For in-hospital mortality prediction, LSTM models and transformer models were trained for 100 epochs using the MIMIC-III and eICU datasets separately. For 5-year cancer survivability prediction, MLP models were trained for 25 epochs with the SEER BCS or LCS dataset with optimal hyperparameter settings mentioned^19^. Other models, including XGBoost, AdaBoost, and Random forest, are trained using the best hyperparameters obtained from grid search (Supplementary Table 1). The models were trained using binary cross-entropy loss. An epoch was selected based on the threshold-agnostic validation area under the precision-recall curve (AUPRC) and validation loss to avoid overfitting. Specifically, we first selected the top 3 epochs with the highest validation AUPRC and then selected the epoch with the minimum validation loss (Supplementary Tables 2 and 3). We monitored the validation loss and training loss difference to prevent overfitting. In all experiments, the chosen machine learning model demonstrated a small loss difference (Supplementary Tables 2 and 3).

Besides evaluating models trained on the original training sets, we also experimented with resampling and reweighting techniques and measured how well the resulting bias-corrected machine learning models performed in our critical zone tests. The reweighting technique has demonstrated superior performance in healthcare datasets, as evidenced by prior studies^24^. For resampling, we tested two generative resampling approaches, SMOTE (Synthetic Minority Oversampling Technique) and AdaSyn (Adaptive Synthetic Sampling). We employed Python’s Imblearn library to apply SMOTE and AdaSyn oversampling techniques, generating balanced training sets by increasing samples from the minority class (sizes shown in Supplementary Table 4). For reweighting, we utilized Python’s Sklearn library to compute balanced class weights based on the training sets (Supplementary Table 5). These methods are applied to the LSTM model for mortality prediction and to the MLP model for cancer survivability prediction.

The training, validation, and test set breakdown for MIMIC-III and eICU datasets is 70%, 15%, and 15% and 80%, 10%, and 10% for the BCS and LCS datasets. After model calibration, a threshold-tuning process is conducted on the validation set, and an optimal threshold is selected based on balanced accuracy and F1 score for the minority class. Specifically, after training, we first conducted model calibration by applying Isotonic Regression using the validation set. Model calibration mapped the predicted probabilities to actual probabilities. Then, we performed threshold tuning to determine the optimal threshold. The minority F1 score and balanced accuracy were computed on the validation set for each threshold ranging from 0.0 to 1.0 with a step size of 0.01. Subsequently, the top three thresholds yielding the highest minority F1 scores were identified, and the optimal threshold maximizing balanced accuracy across all validation samples was selected. This process was repeated for 3 independently trained models of each type, and the average threshold was calculated from these independent trials. Thresholds are shown in Supplementary Table 6. The tasks were executed on a machine with Ubuntu 18.04 operating system, x86-64 core-i9 architecture, 8 physical cores (16 virtual cores), and 32 GB RAM. The experimental code and models were written using Python 3.7, TensorFlow 1.15, and Keras 2.1.2. The cancer survivability prediction MLP model was trained on a machine with x86_64 Intel(R) Xeon(R) CPU 2.40GHz (40 cores) and 125 GB RAM. The experimental code and model were written using Python 3.6, TensorFlow 2.9.0, and Keras 2.9.0.

### Mapping neuron activations

We visualized the activated neurons in a neural network model for a particular input. The Keras backend was used to capture the neuron outputs from the bidirectional layer output and LSTM layer output for the mortality risk prediction model. Sigmoid activation was applied to obtain neuron output values in the range of [0, 1]. To quantify changes in neuron activation, we defined and computed Neural Zone Activation (NZA) and average zone difference Δ*NAZ*. A zone is defined by the attribute range bounded by two values. NZA calculates the average neurons’ activations within a zone, where a zone can be a critically low, critically high, or normal range (Supplementary Equation 1). Δ*NAZ* computes the average NZA difference between two zones (Supplementary Equation 2), such as normal and critically high zones, indicating how much neurons react to zone changes. There is no standard value for Δ*NAZ*. A relatively higher value indicates a good response.

### Statistical methods

Model performance is reported using the average and standard deviations, which are calculated using 9 or 15 trials. The trials were performed using 3 model instances that have identical architecture and were trained on the same training set with random model parameter initialization. Each of the 3 model instances is evaluated with 3-5 test sets. The distribution shift of the synthesized test dataset from the original training sets was quantified by Wasserstein distance (WD)^25,26^. We used an implementation from the Python library called scipy.stats.wasserstein_distance. First, the WD was calculated between the same features from the whole original dataset and the synthesized test set. Then, the feature-specific WD was averaged to obtain the mean WD for quantifying the distribution shift.

### Attribute-based test case generation for in-hospital mortality risk prediction

We created new cases by increasing or decreasing one or multiple vital health parameters in the seeding records. To reduce computing complexity, we prioritized by focusing on the most influential features. Relevant medical terminologies are explained in the Supplementary Notes.

In the *single-attribute variation*, *w*e generated new test cases by varying a single attribute at a time while keeping other attributes unchanged. We then evaluated how the model reacts to these changes and its ability to recognize associated risks (e.g., hypoglycemia). Specifically, given an attribute *A*, single-attribute variation for time series involved the following operations. First, we identified *A*’s minimum and maximum values in the MIMIC-III or eICU datasets, which defined the observed range.

Then, the mean and the variance of attribute *A* were computed from the entire dataset. Using the variance and the observed range, we generated a series of random values for every value from that range, one value for each of the 48 hours. Then, the new test case was formed by having these generated values for attribute *A* and other attribute values directly inherited from the seed. We repeat this process for every possible attribute value from the observed range with step 1.

*Multi-attribute variation* generated new test cases by modifying two or more attributes, aiming to represent medical conditions that were characterized by variations in multiple related attributes. We further differentiated two scenarios: *i)* a single set of medically correlated attributes driven by one underlying disease condition, e.g., high diastolic and systolic blood pressure due to hypertension, and *ii)* medically correlated attributes due to multiple underlying conditions, e.g., hypertension and diabetes.

These test cases were used to assess the machine learning model’s ability to respond to the risks of multiple disease conditions in patients. One of the test sets was created by changing multiple vitals such as systolic blood pressure, diastolic blood pressure, blood glucose level, respiratory rate, heart rate, and body temperature at the same time. A test case was assigned a ground truth label using existing literature or under the guidance of medical doctors. 6 multi-attribute test cases and 12 deteriorating test cases were directly labeled by the medical doctor (Supplementary Tables 7 and 8).

### Deteriorating test case generation for MIMIC-III

We leveraged the gradients of LSTM to guide the generation of new test cases. This method is automatic and does not require the specification of attributes to change, aiming to generate new test cases that are challenging for machine learning models to classify correctly. Such cases typically occur at the decision boundary of the classifier. Our method started from a healthy patient’s record (i.e., a seed with low or zero mortality risk). The seed is a time-series record far away from the classifier’s decision boundary. We incrementally adjusted the attribute values of the seed by following the steepest direction (i.e., gradient) that can *maximize* the loss (i.e., prediction errors of the machine learning model). This process explores the local hyperspace and iteratively produces new cases that are closer and closer to the machine learning model’s decision boundary. Computationally, given a trained machine learning model and a healthy patient’s time series record as the seed, we computed the derivative of the model’s loss function, i.e., gradient (Supplementary Equation 3). The gradient is a vector of partial derivatives describing the direction and rate of changes of the loss function. Then, we changed the test case in the direction of increasing gradient. Our algorithm is described in the Supplementary Methods Section. The step size or learning rate to control the magnitude of the change was set to 0.001-0.2 in our experiment depending on the attribute (Supplementary Table 9). Our method focuses on generating samples; it differs from the common gradient descent process, which adjusts model weights to minimize loss. We have two ways of creating gradient-based test cases from the MIMIC-III dataset -- *single-attribute gradient approach* and *multi-attribute gradient approach*. In the former, we focus on a single attribute and apply gradient ascent solely to modify that specific attribute. This approach allows one to observe the individual impact of each attribute on the mortality risk. In the latter, we simultaneously change values of multiple attributes using gradient ascent. Gradient approaches create test cases that represent deteriorating health conditions in continuous time series. Supplementary Table 10 shows the various categories of test sets and their sizes.

### Glasgow coma scale test case generation

The Glasgow Coma Scale (GCS)^27^ is a neurological scale that assesses a patient’s level of consciousness. It evaluates responses in three categories: eye-opening (E), verbal response (V), and motor response (M), adding up to a score ranging from 3 to 15. A lower score indicates a more severe impairment of consciousness. Definitions of values in each category are given in Supplementary Table 11. A GCS score can be representative of multiple sets. For example, GCS total 10 can be the outcome of (E, V, M) = (3, 3, 4), or (4, 4, 2), etc. The GCS total test set contains all the possible combinations of (E, V, M) for each particular GCS total value. The double attribute-based GCS cases were also created by varying both attributes and keeping the other constants to healthy values.

### Attribute-based test case generation for 5-year cancer prognosis

#### Single-attribute variation

Similarly, we engineered cancer test cases by varying one attribute of a seed record. The attribute may be the size of the tumor (T), the number of positive lymph nodes (N), the number of examined lymph nodes (ELNs), or the grade of the cancer cell. T and N are the two most important factors for determining cancer severity or stage^28^. T has 4 categories based on the size. The tumor test set was created by varying the size of 3 seeds in the surviving class, using the range (0-986 mm) from the original SEER dataset. This BCS tumor size test set contains 12,891 cases, including 18 T0 cases, 243 T1 cases, 390 T2 cases, and the rest of 12,240 T3 cases. The LCS tumor size contains 8,367 cases, including 12 T0 cases, 171 T1 cases, 273 T2 cases, and 7,911 T3 cases. (T4 cases cannot be created, as it is not associated with a quantitative value). The number of positive lymph nodes (N) is divided into 4 categories. The positive lymph node test case was created similarly by changing the corresponding value from the same 3 seeds using the attribute range (0-84). For BCS, we generated 7,686 test cases, including 90 N0 cases, 270 N1 cases, 546 N2 cases, and 6,780 N3 cases. For LCS, we generated 24,264 test cases, including 333 N0 cases, 999 N1 cases, 1,998 N2 cases, and 20,934 N3 cases. Supplementary Notes have more details of T and N category definitions.

The ELN test case was created similarly by varying the number of ELNs (range in [0, 86]) from 3 seeds and keeping other values the same as the seeds. The ELN test set contains a total of 3,510 cases for BCS and 1,835 cases for LCS. Although the number of examined lymph nodes (ELNs) is not directly related to the cancer staging, it is crucial for diagnosing cancer. Several studies proposed that there should be a standard (or a minimum) number of ELN cancer diagnoses^29–32^. The grade of the cancer cell represents the spreading and growth intensity of the cancer cell^28^. The SEER dataset contains 1-4 grades where the higher grade represents faster growth and speed and another grade 9 for undetermined (not stated/applicable). For BCS, we created test sets for each of 1-4 grades, where each set contains 24,875, created from 21,723 cases from the majority Class 1 (survival) and 3,152 cases from the minority Class 0 (death). We utilized the entire validation set as the seed pool, allowing for a more comprehensive evaluation. In total, the 1-4 grade test set contains 99,500 cases (24,875 cases for each grade). To create each grade test set, we set the corresponding grade value to all data points in the validation set.

#### Double- and triple-attribute variations

Double-attribute variation generated new BCS test cases by changing a pair of attributes from the 3 continuous attributes, *i)* the size of the tumor (T), *ii)* the number of positive lymph nodes (N), and *iii)* the number of examined lymph nodes (ELNs). The grade attribute was excluded, as it is categorical. The tumor size and positive lymph node combination test set contains 18,531 test cases. The tumor size and number of examined lymph nodes combination test set also contains 18,531 test cases. The number of examined lymph node and positive lymph node combination test sets contain 23,400 test cases. The triple-attribute test set was created by setting three attributes simultaneously to represent serious disease conditions, e.g., tumor size to T4, number of positive lymph nodes to N3, and grade to 4. The validation set, consisting of 24,875 cases including 21,723 cases from Class 1 (survived) and 3,152 from Class 0 (death), was used as seeds. While the tumor size and number of positive lymph nodes are continuous variables, we treated them as categorical by selecting a value from the T4 and N3 range respectively. As a result, the triple-attribute test set contains 21,723 cases derived from Class 1 seeds and 3,152 cases derived from Class 0 seeds. Supplementary Table 12 summarizes the various categories of test sets and their sizes. We performed double- and triple-attribute variation tests for BCS models, not on LCS models.

For labeling generated breast and lung cancer test cases, we used authoritative literature to assign labels. We labeled cases with Class 0 (indicating low survivability) if there was a strong presence of cancer (i.e., T 1-3, N 1-3, and grade 2-4). For ELNs, the previous studies using SEER datasets^33,34^ suggested using at least 8-9 ELNs for stage T1 diagnosis, 37 ELNs for T2 diagnosis, and 87 ELNs for T3 diagnosis. As ELN is not directly responsible for the death, that attribute was not considered during labeling.

#### Selection of Seeds

We used existing patient records from the original dataset as seeds (i.e., starting points) to generate synthetic test sets. We selected seeds from the in-hospital mortality dataset that are real-world non-death patient cases that exhibit healthy attribute values. Seeds were chosen as follows. For attribute-based test case generation, we randomly selected seeds from MIMIC-III Class 0 (survival case) following two criteria. First, the mean (of 48 hours) attribute values are within the range of ideal health conditions defined in Supplementary Table 13. In addition, the standard deviation of each attribute needs to be less than or equal to the mean standard deviation (Supplementary Table 14) of the MIMIC-III dataset. Our evaluation of attribute-based test case generation involved 5 seeds and the statistics of these 5 seeds are given in Supplementary Table 15. The deterioration test case generation involved another 3 seeds, which were selected randomly from Class 0 of MIMIC-III. Since the eICU dataset contains similar samples with identical features and a consistent 48-hour time duration, we utilized the same test set generated from MIMIC to evaluate models trained on the eICU dataset. Additionally, the selected seed attributes fall within the healthy (ideal) range, minimizing the out-of-distribution effects on models trained on the eICU dataset. For the cancer survivability prediction task, test cases involving changing a numerical variable were generated from 3 randomly selected seeds from the surviving class. Test sets are separately generated from each of the SEER BCS and LCS datasets. Test sets involving categorical variables, such as grades test and triple-attribute test sets, were generated using all validation data points from SEER.

## Results

For in-hospital mortality prediction, we generated 177,507 new time-series test cases based on MIMIC-III to represent serious patient conditions and used them to evaluate the responsiveness of machine-learning models (Supplementary Table 10). 126,950 cases are generated by modifying multiple vital attribute values in 5 seed records, 42,500 cases by modifying double attributes in seed record, 7,075 cases by modifying a single attribute value in a seed record, 970 cases by modifying Glasgow coma scale (GCS), and 12 cases by gradient ascent. Modifications to vital attributes are bounded by the minimum and maximum values of the attribute in the in-hospital mortality (IHM) datasets and focus on critically high and critically low ranges of the 6 vitals. We carefully use literature^35–39^ to identify these ranges (Supplementary Table 13). The test case generation also ensures the continuity of the time series. The 6 types of attributes include systolic blood pressure, diastolic blood pressure, blood glucose level, respiratory rate, heart rate, and body temperature. A seed record is a real-world patient case selected from the MIMIC-III dataset that is a non-death case whose attributes are in the typical healthy ranges^36–39^. The other 12 test cases are generated using a gradient-ascent approach, which modifies the seed by following the direction of the steepest increasing loss function.

Each synthetic test case is assigned a label, death (Class 1) or survival (Class 0) for in-hospital mortality (IHM) prediction. Labels are verified either by a medical doctor or confirmed by the literature. These labels are considered ground truth in our study. Two medical doctors reviewed 18 generated test cases (6 attribute-based cases and 12 gradient-based cases), where the test cases are time series data and the risk scores of the medical doctors’ output are quantitative, between 0 and 1. The medical experts estimated risk values are in Supplementary Tables 7 and 8. Labels of the other 177,489 test cases are inferred based on expected ranges of vital health parameters of healthy individuals extracted from medical literature.

Synthetic test cases persistently containing vital values in critical zones represent patients in sustained critical health conditions, and thus are labeled Class 1. These cases should receive a high mortality risk prediction from machine learning models. We define *machine learning responsiveness* as the model’s ability to react to substantial changes in input values, e.g., by increasing the mortality risk score for IHM prediction.

Based on the SEER 5-year breast cancer survivability dataset, we generated 205,414 test cases to represent different patient conditions. Among them, 120,077 cases are generated by changing single attributes (including 7,686 cases representing different N stages, 12,891 cases for the T stage, and 99,500 cases for grades), 60,462 cases by modifying double attributes, and 24,875 cases by changing triple attributes from the seed cases. Based on the LCS dataset, we generated three sets of single-attribute test cases totaling 31,136 cases, which include 8,367 cases representing different T stages, 24,264 cases representing N stages, and 1,835 cases representing ELNs (Supplementary Table 12). We manually assigned labels to synthesized test cases guided by the literature^28–34^.

### ML performance under Glasgow Coma Scale (GCS) testing

For in-hospital mortality prediction, we assess MIMIC III-based LSTM, CW-LSTM, and LR models with test cases containing varying GCS scores (Figure 2), including severe injury cases with GCS scores 3 to 8, moderate injury with 9 to 12, and mild or no injury with 13 to 15. A low GCS score indicates a poor health condition^27^ (medical meanings of each category are shown in Supplementary Table 11). The CW-LSTM model gives near zero mortality risk values for 15 severe injury cases, for example, E4M1V3 in Figure 2a, i.e., a case with an eye response score of 4 out of 4, a motor response of 1 out of 6, and a verbal response score of 3 out of 5. For a moderate injury case E4M1V5, CW-LSTM also gives an unexpectedly low mortality risk (0.01) prediction, i.e., predicting the healthy outcome of the patient. The model’s prediction is inconsistent, as another moderate injury case E4M3V5 receives a high mortality risk of 0.58.

**Figure 2:**
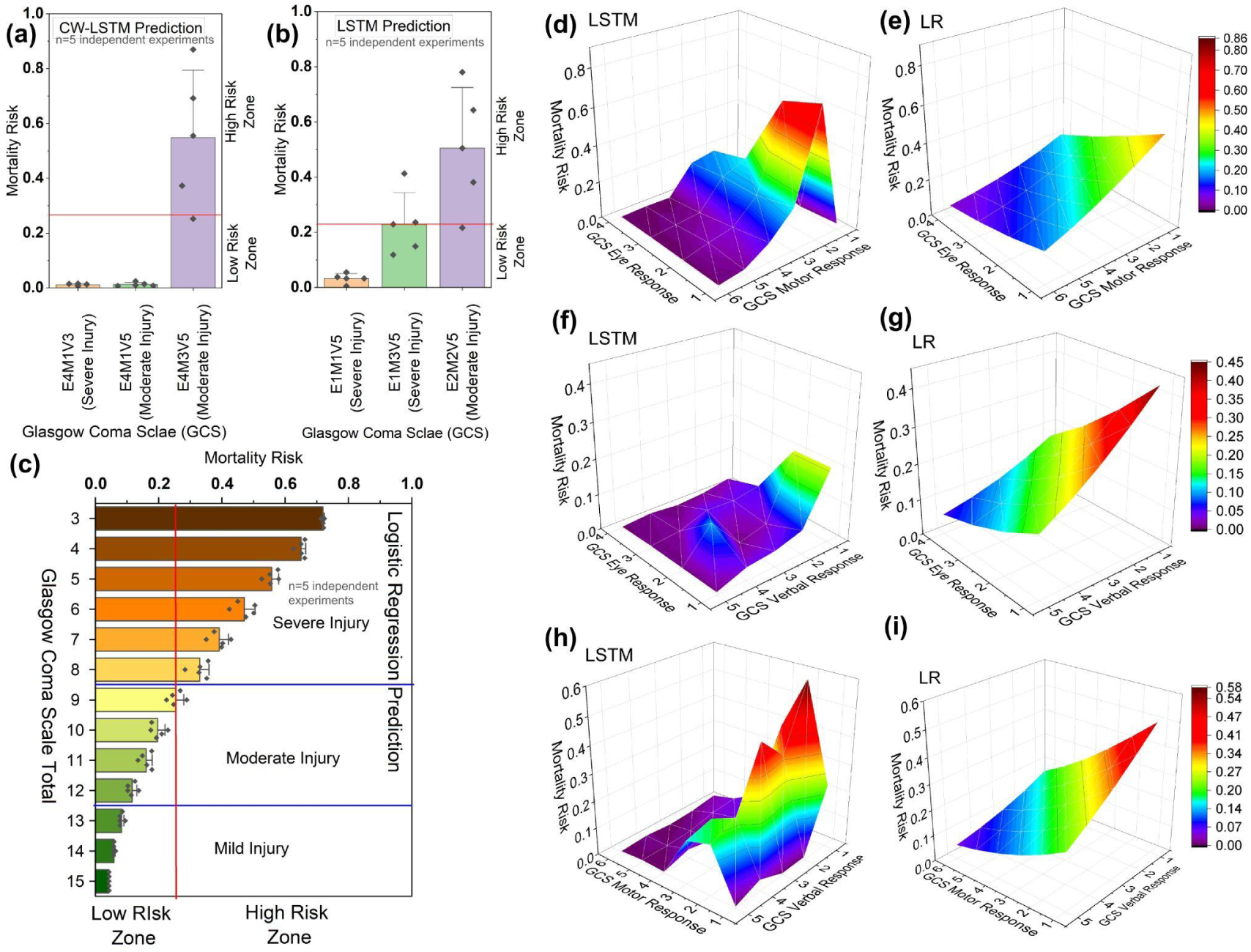
Mortality risk (MR) prediction for Glasgow Coma Scale for different combinations using three machine learning models. MR predicted by (a) channel-wise LSTM model for three injury cases, (b) LSTM model for three injury cases, and (c) Logistic regression for injury cases indicated by all combinations of GCS scores. MR prediction of injury cases defined by different combinations of GCS eye and motor response scores by (d) LSTM and (e) logistic regression model. MR predicted by (f) LSTM and (g) logistic regression using injury cases defined by different combinations of GCS eye and verbal response scores. MR prediction of injury cases defined by different combinations of GCS motor and motor response scores by (h) LSTM and (i) logistic regression.

Similar inaccuracies and inconsistencies are also observed for the LSTM (MIMIC III) model tested. For instance, the LSTM model mistakenly considers a severe injury case E1M1V5 to be much more likely to survive than a moderate injury case E2M2V5 (Figure 2b). In contrast, the LR model consistently predicts at least 0.3 mortality risk for severe injury cases and responds well (Figure 2c). For mild injury cases, the LR model consistently predicts a low mortality risk. The 3D surfaces of the LR model appear smooth and the model reacts to decreased eye and motor signals (Figure 2e). In contrast, LSTM’s 3D plots are less monotonic, exhibiting bumps (Figure 2). For the most severe cases (subscores being 1 or 2), LSTM’s risk predictions incorrectly drop.

### ML performance under critical zone tests

#### Single-attribute Critical Zone Test Results

We evaluate the MIMIC III-based LSTM, CW-LSTM, and LR models’ ability to respond to a single deteriorating attribute while keeping other attributes stable as in the seed (Figure 3). The CW-LSTM model fails to recognize bradypnea, i.e., an abnormally slow breathing rate and gives only slightly elevated mortality risk prediction (mean mortality risk 0.05 and standard deviation 0.04) for tachypnea, i.e., rapid breathing (Figure 3a), insufficient to trigger an alert. Similarly, CW-LSTM is unable to recognize most of the abnormal vitals. Its mortality risk prediction gives a negligible change to an abnormal patient’s glucose level (Figure 3c) and oxygen saturation rate (Figure 3f). For the other 3 attributes tested, CW-LSTM gives small partial responses to either a high critical zone or a low critical zone, but not both. For example, it is unable to recognize high body temperature anomalies and only slightly raises the mortality risk to 0.01 to 0.08 (standard deviation 0.033) for severe hypothermia below 34°C (Figure 3b), which is still much below the classification threshold (0.22). CW-LSTM’s response to abnormal diastolic and systolic blood pressure is also inadequate (Figures 3d and 3e).

**Figure 3:**
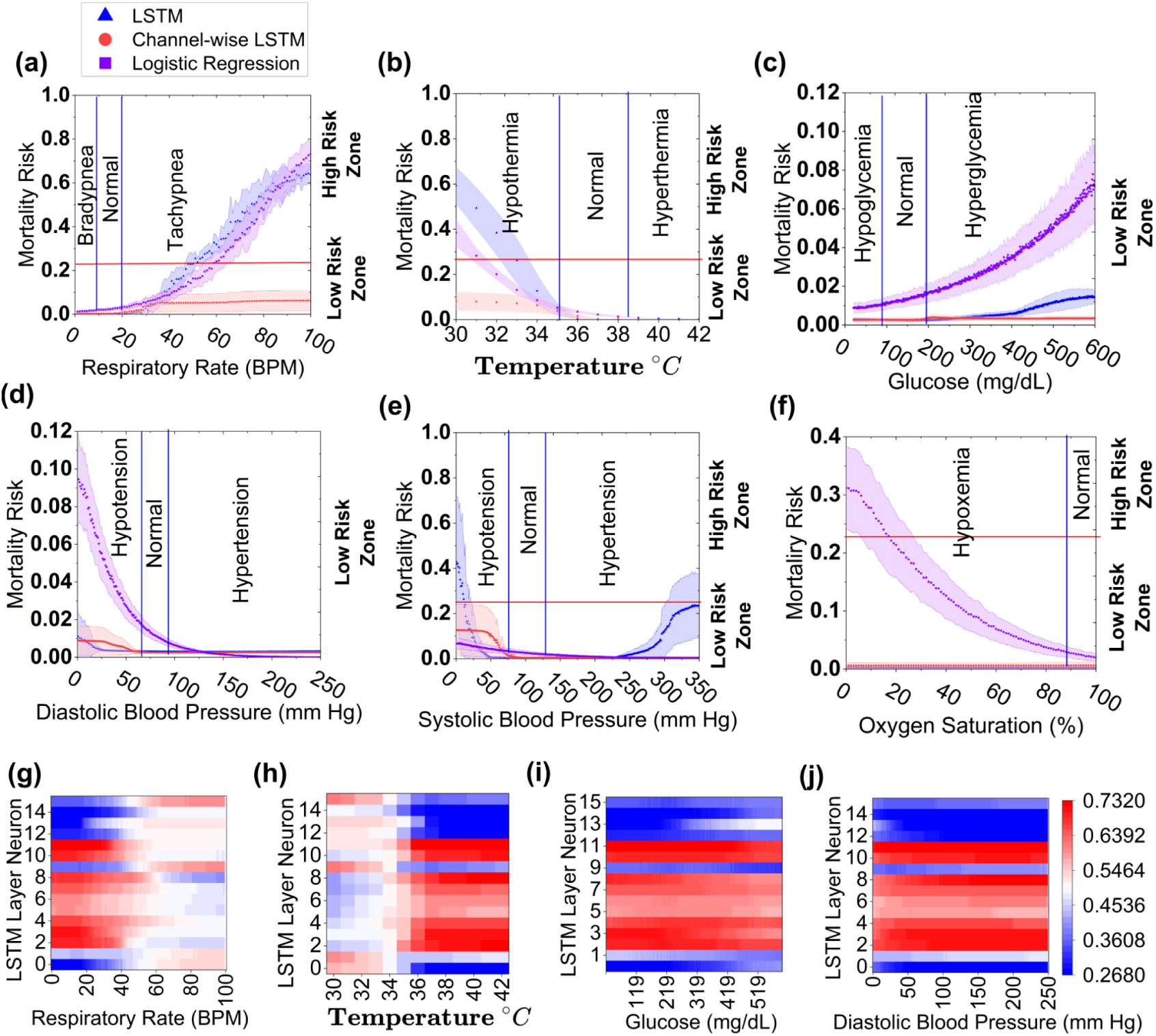
Mortality risk prediction for single vital-sign tests using three machine learning models (LSTM, Channel-wise LSTM, and Logistic Regression) and visualizing the Neural activation map of LSTM layer consisting of 16 neurons. LSTM, Channel-wise LSTM, and Logistic Regression (LR), predict the mortality risk (MR) of (a) respiratory rate, (b) body temperature, (c) glucose, (d) diastolic blood pressure, (e) systolic blood pressure, and (f) oxygen saturation test sets (synthesized). The mortality risk (MR) is represented by X-axis and MR above and below a red horizontal line (threshold = 0.22) indicates a high or low mortality risk zone respectively. The entire range of each vital sign (except oxygen saturation) value is divided into three segments, low, normal, and high, by the blue vertical lines. The low and high values within these ranges indicate critical health conditions. Figures in the last row (g)-(j) represent the neural activation map. These are the neural activation values, calculated after applying the sigmoid function, when the model is fed with test cases varying a single vital, such as (g) glucose, (h) diastolic blood pressure, (i) temperature, and (j) respiratory rate.

The LSTM model gives much more elevated risk prediction than CW-LSTM for tachypnea (Figure 3a) and hypothermia conditions (Figure 3b). Out of the 3 models tested, LSTM is the only machine-learning model that responds to both systolic hypotension and hypertension conditions, producing a U-shaped curve (Figure 3e). However, LSTM consistently gives an ultra-low risk prediction for abnormal diastolic blood pressure (Figure 3d). Similar to CW-LSTM, LSTM does not recognize hypoxemia, i.e., low blood oxygen level (Figure 3f), bradypnea, hyperthermia, and abnormal glucose level (Figure 3c), exhibiting either monotonic or near-flat risk prediction curves insensitive to abnormal vitals. Compared to the other models, logistic regression gives a substantially higher risk prediction for hypoxemia. It also computes elevated risk scores in response to increasing hyperglycemia and diastolic hypotension conditions. For all attributes, logistic regression is only able to recognize one end of the critical zones, but not both. Overall, logistic regression, LSTM, and CW-LSTM correctly predict 37.7%, 37.8%, and 22.4% of the single-attribute critical zone test cases on average (Supplementary Table 16).

### Neuron activation analysis

We visualized neuron output from intermediate layers of the MIMIC III-based LSTM model. Neurons whose activations change with changing variable values are the responsible neurons for recognizing that variable. We found most of the LSTM neurons have low or no responses to varying glucose and diastolic blood pressure values (Figures 3i and 3j). In contrast, neurons are more responsive to temperature and respiratory rate changes, e.g., sharp changes in all neuron activation between 34°C to 36°C (Figure 3h) and around or above 40 bpm (Figure 3g). However, neurons exhibit minimal or no changes in activation for higher temperatures or for critically low respiration rates.

To quantify changes in neuron activation, we computed Neural Zone Activation (NZA) and average zone difference Δ*NAZ*, new metrics defined by us (Supplementary Equations 1 and 2). NZA averages neurons’ activations within a zone, where a zone is critically low, critically high, or normal range (Supplementary Equation 1). Δ*NAZ* computes the averaged NZA difference between zones (Supplementary Equation 2), indicating how much neurons react to zone changes. The LSTM model shows low (0.01 to 0.04) Δ*NAZ* in most cases (Supplementary Table 17). In a few cases, e.g., temperature Δ*NAZ* (low, normal) and respiratory rate Δ*NAZ* (high, normal), the values are relatively high (0.14 to 0.16).

#### Multi-attribute Critical Zone Test Results

We evaluated the 3 MIMIC III-based machine learning models under 42,500 double attribute varying test cases (Figure 4), including respiratory rate and heart rate pair (first row), systolic and diastolic blood pressure pair (middle row), and glucose and diastolic blood pressure pair (last row). The CW-LSTM model does not generate high mortality risk predictions for most critical zone cases, consistent with its single-attribute test performance in Figure 3. The logistic regression model gives better performance than CW-LSTM, predicting higher risks for some critical zone combinations (Figures 4a, 4d, and 4g).

**Figure 4:**
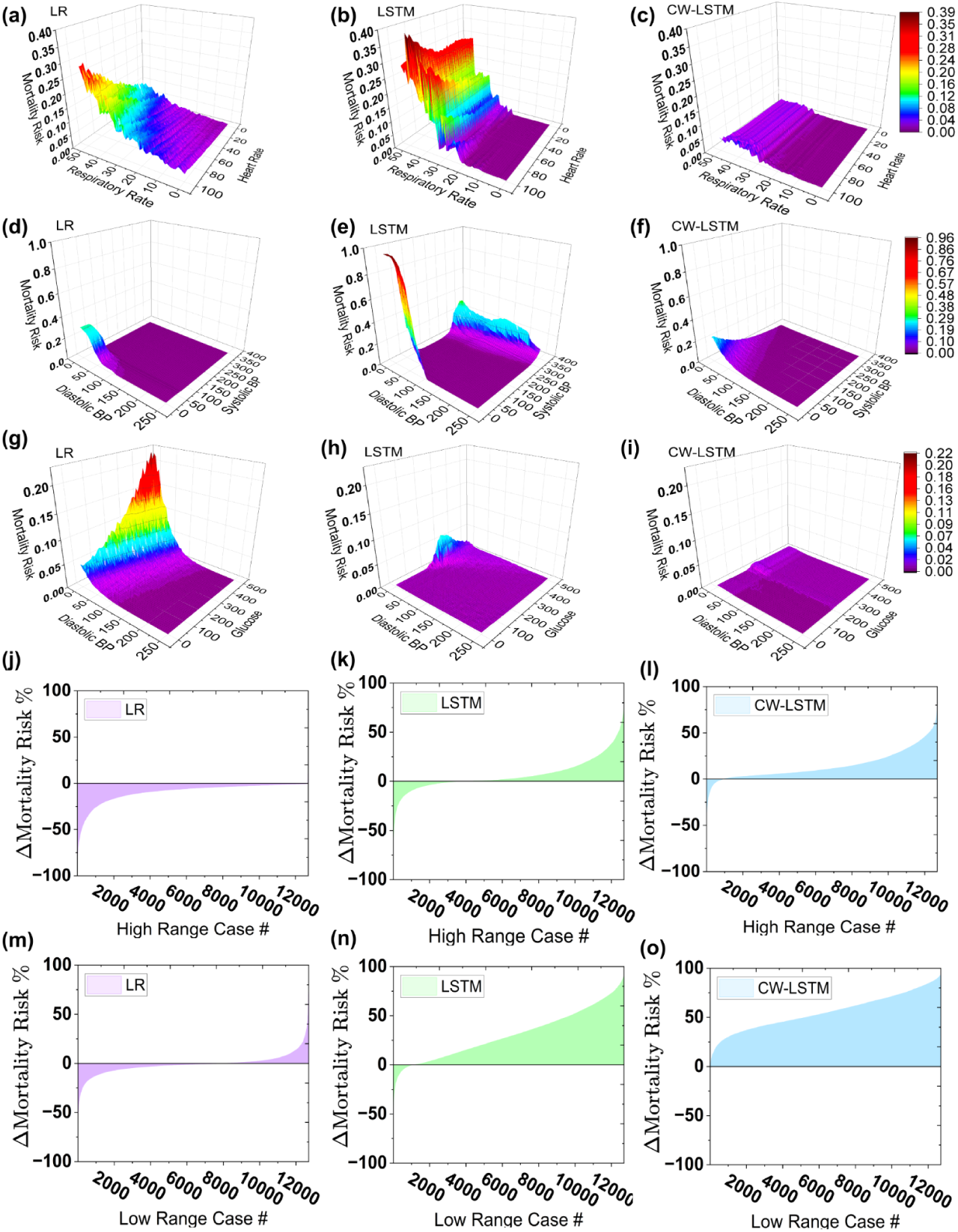
(a)-(i) show the mortality risk prediction of patients attributed by double vital-sign and (j)-(o) show the mortality risk difference (ΔMR) of seed (Class 0) and test pair generated by altering 6 attributes at the same time. Risk prediction under varying respiratory rate and heart rate by (a) logistic regression, (b) LSTM model, and (c) CW-LSTM model. Risk prediction under varying systolic and diastolic blood pressure by (d) logistic regression, (e) LSTM model, and (f) CW-LSTM model. Risk prediction under varying glucose and diastolic blood pressure by (g) logistic regression, (h) LSTM model, and (i) CW-LSTM model. (j), (k), and (l) represent ΔMR for high critical range cases and (m), (n), and (o) represent ΔMR for low critical range cases. The test set is generated by simultaneously varying systolic blood pressure, diastolic blood pressure, blood glucose level, respiratory rate, heart rate, and body temperature and values are randomly selected from the critical zone. The graph shows mortality risk difference (ΔMR) calculated by subtracting predicted mortality risk of the seed data from the predicted mortality risk of its corresponding critical case. The X-axis represents the case numbers and the Y-axis represents ΔMR. It is expected to get a positive MR difference and the negative ΔMR cases represent failed test cases.

However, its prediction is monotonic, thus, unable to recognize both high and low critical zones of an attribute pair. For example, logistic regression fails to alert when patients have low respiratory rate and low heart rate. LSTM model exhibits prediction behaviors (Figures 4b, 4e, and 4h) consistent with its single attribute performances in Figure 3.

In a 6-attribute varying test setting, we evaluated the responsiveness of MIMIC III-based machine learning models under 6 changing vitals, where test cases have abnormal systolic and diastolic blood pressures, blood glucose level, respiratory rate, heart rate, and body temperature values in their respective critical zones. We recorded how much mortality risk scores changed and showed the distributions in Figures 4j to 4o. Figures 4j, 4k, and 4l show the mortality risk difference (ΔMR) between each high critical zone test case and its corresponding seed. Medically speaking, the risk should increase under worse health conditions. The CW-LSTM model consistently predicts high mortality risk for most cases, resulting in a positive ΔMR for over 90% of the cases (Figures 4i and 4o). The logistic regression model (Figure 4j) produces negative ΔMR for all cases, which is incorrect. The LSTM model generates positive ΔMR for two-thirds of the 12,694 test cases.

The 3 models under low critical zone tests performed similarly (Figures 4m, 4n, and 4o), where CW-LSTM responds to multi-attribute critical conditions the most effectively and logistic regression the least. Overall, logistic regression, LSTM, and CW-LSTM correctly predict 6.2%, 45.7%, and 69.3% of the multi-attribute critical zone test cases on average (Supplementary Table 16), respectively.

### Results on test cases with deteriorating conditions

For in-hospital mortality prediction (using the MIMIC III dataset), we used a gradient ascent method to generate 12 time-series test cases with deteriorating health conditions. 9 of the 12 test cases contain one vital that worsens during the 48 hours and is in the critical zone during the last 24 to 48 hours, including 3 cases of decreasing systolic blood pressure (Figure 5a), 3 cases of increasing respiratory rate cases (Figure 5b), and 3 cases of decreasing body temperature (Figure 5c). The other 3 test cases have multiple (3) worsening vital signs, with vitals being in critical zones during the last hours (ranging from 30 to 48 hours). All 12 test cases should receive a high mortality risk prediction, i.e., Class 1. We confirmed these labels with two medical doctors who manually reviewed the time series data.

**Figure 5:**
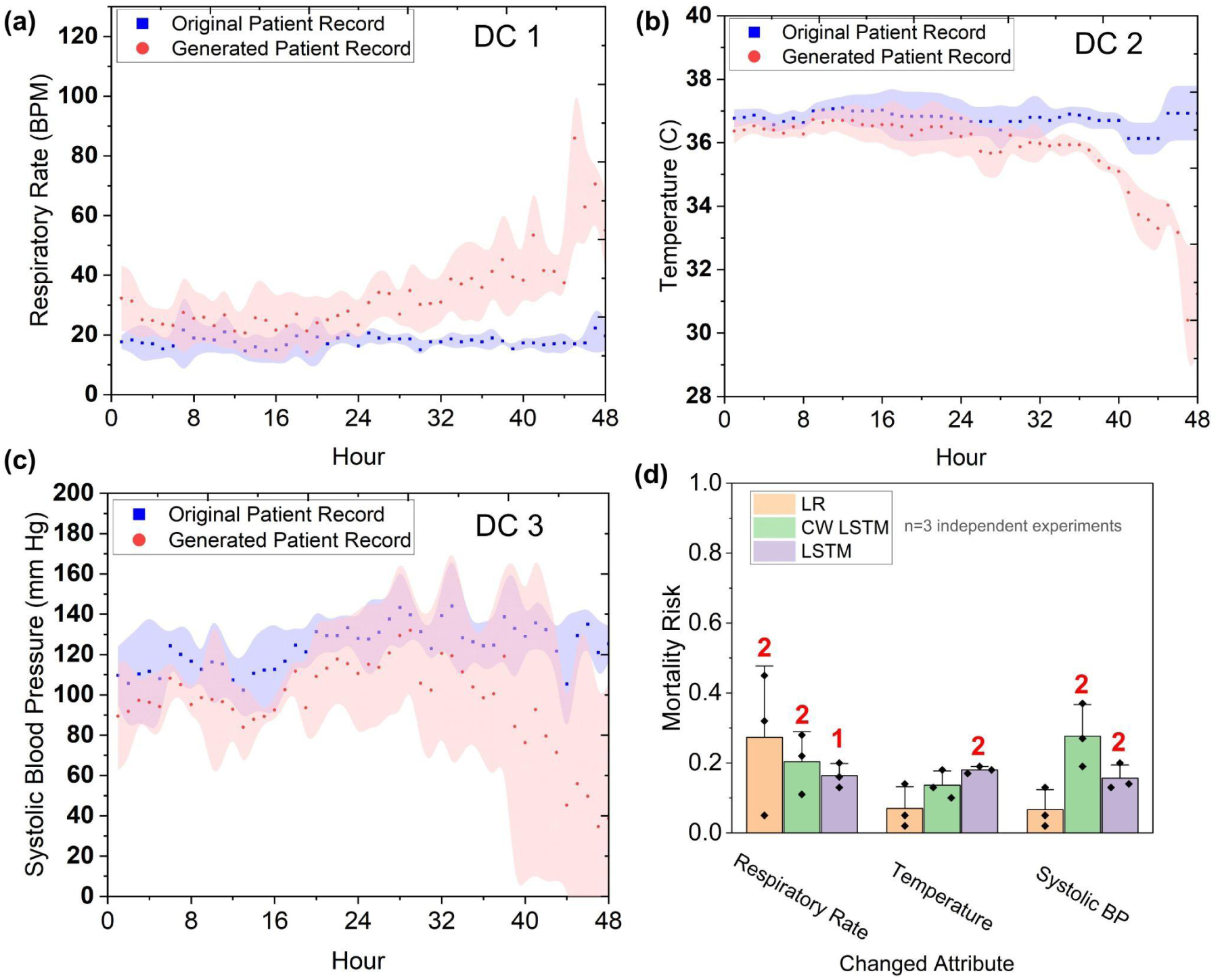
Gradient-generated deteriorating test cases and machine learning models’ mortality risk predictions by LR, CW-LSTM, and LSTM models. (a)-(c) show the average time series of the generated abnormal test cases (in red area curves) and the normal seed cases used (in blue area curves) for each of the 3 attributes. (d) Models’ predicted average mortality risks for each deteriorating attribute. The standard deviation is indicated by the error bar. The numbers (red) at each of the bars represent the number of detected alerts out of input 3 cases.

Average mortality risks predicted by machine learning models on the 9 single-attribute deteriorating test cases are in Figure 5d. Out of the 9 single-attribute deteriorating test cases, logistic regression only detects 2 (22%) respiratory rate cases (average risk 0.38) and fails to detect the other 7. CW-LSTM (MIMIC III) detects 4 (44%) out of the 9 deteriorating test cases, including 2 systolic BP cases (average risk 0.32) and 2 respiratory rate cases (average risk 0.39). However, it is unable to detect deteriorating temperature cases. LSTM reports 5 (56%) out of the 9 deteriorating test cases, including 2 systolic BP cases (average risk 0.22), 2 temperature cases (average risk 0.23), and 1 respiratory rate case (risk 0.22). Collectively, the models detect 41% out of single-attribute deteriorating test cases.

Multi-attribute test cases have 3 deteriorating vitals, including oxygen saturation, temperature, and diastolic blood pressure. The LSTM (MIMIC III) model detects all 3 cases (average risk 0.23), whereas CW-LSTM fails to generate any alerts. Logistic regression (MIMIC III) issues alerts for 2 out of 3 cases (average risk 0.56). Collectively, the 3 models detect 56% out of the multi-attribute deteriorating test cases. Overall, the models’ average accuracy under all deteriorating test cases is 44%.

### 5-year cancer survivability results

We found similar deficiencies in the machine learning model, in terms of the model’s ability to respond to test cases representing serious cancer conditions.

#### Single-attribute test results

We evaluated the responsiveness of MLP models trained on the BCS dataset to a single deteriorating attribute (Figure 6 and Supplementary Table 18) while keeping other attributes the same as the seed. The BCS-MLP model shows some responsiveness with varying tumor size (Figure 6a), however, remains above the survivability threshold (0.71) in all cases. As a result, the model fails to trigger an alert for tumor sizes representing critical stage T1 (tumor size less than 20 mm) to T3 (tumor size larger than 50 mm). On the other hand, the model triggers alerts for 74.4% of the 6,780 N3 stage (Figure 6b). It fails to generate any alert for 270 N1 and 546 N2 stage cases. The BCS-MLP model accurately generates alerts for 66.4% of critical cases (N1-N3). It decreases the survivability for lower numbers of examined lymph nodes (ELNs), however, still fails to trigger any alerts (Figure 6c).

**Figure 6:**
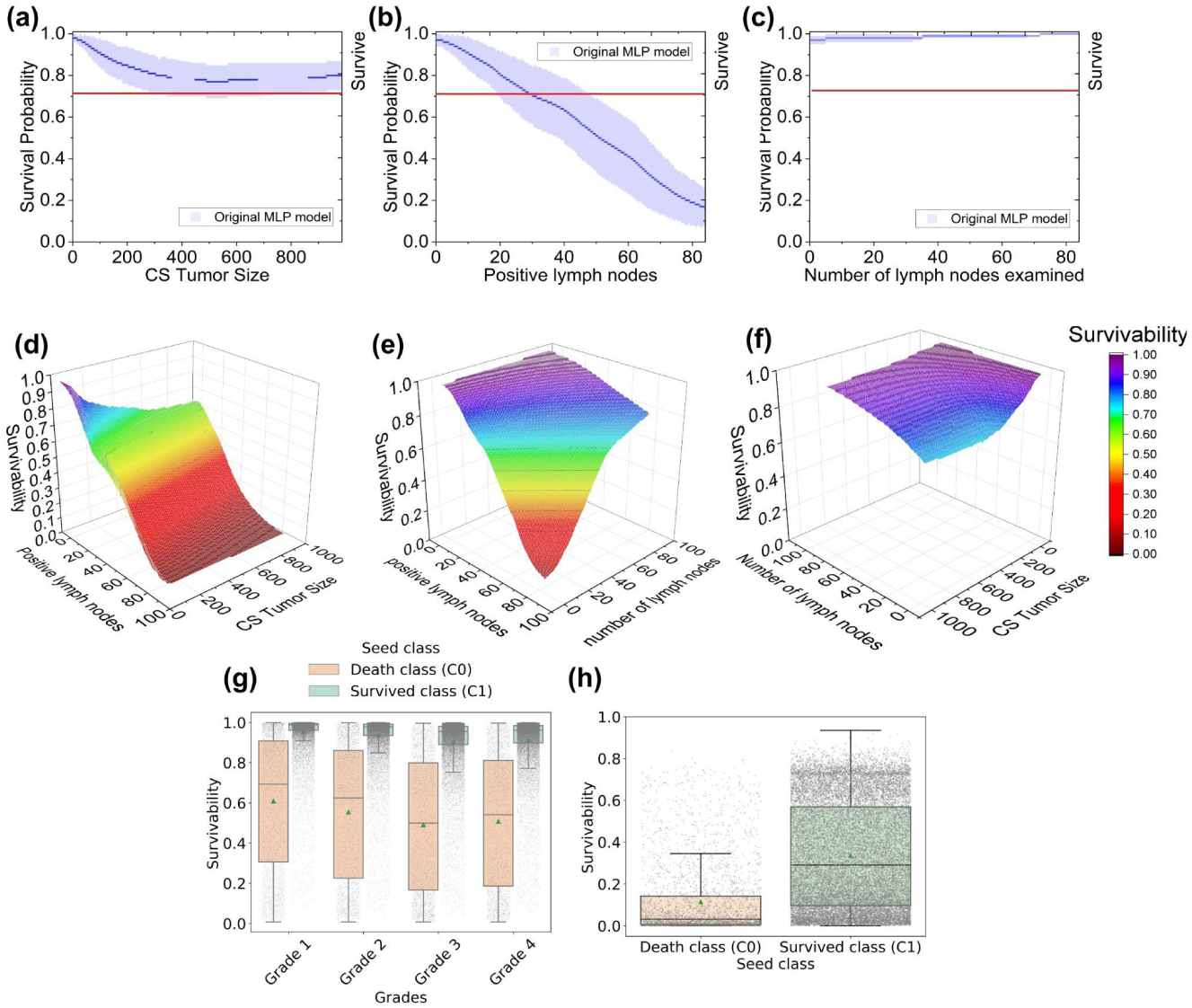
Predicted 5-year breast cancer survivability results of a multi-layer perceptron (MLP) model on test cases. Four major breast cancer screen attributes are involved, including CS tumor size, number of positive lymph nodes, number of lymph nodes examined, and grade. (a)-(c) and (g) Predicted survivability results on single-attribute varying test cases. The blue area of (a)-(c) represents the standard deviation. (d)-(f) Predicted survivability results on double-attribute varying test cases. (h) Predicted survivability results on triple-attribute varying test cases involving CS tumor size, number of positive lymph nodes, and grade. In the boxplots (g) and (h) the horizontal line within the box represents the median value, while the box itself encompasses the interquartile range (IQR), containing the middle 50% of the data. The whiskers extend to the values within 1.5 times the IQR from the box (upper and lower quartiles). The green triangle point on the box represents the mean of the distribution.

For the grade test sets (1-4) generated from 21,723 surviving patient seeds, the BCS-MLP model generates alerts for 989 cases (4.6%) for grade 2 test, 1,616 cases (7.4%) for grade 3 test, and 1,446 cases (6.7%) for grade 4 test (Figure 6g). On the other hand, for the grade test generated from 3,152 death events, the model generates alerts for 1,826 cases (57.9%) for grade 2 test, 2,073 cases (65.8%) for grade 3 test, and 2,013 (63.9%) cases for grade 4 test. The model did not generate any alerts for 97% and 48.7% of grade 1 cases generated from seeds of survived and death events, respectively. The LCS-MLP model shows higher responsiveness with variations in tumor size (Figure 7j), generating alerts in 80.1% of the test cases (Supplementary Figure S3). It also responds well to varying positive lymph node numbers (Figure 7k) with alerts generated in 92.9% of the cases. However, the LCS-BCS model does not react to the increasing number of lymph nodes examined (Figure 7l).

**Figure 7:**
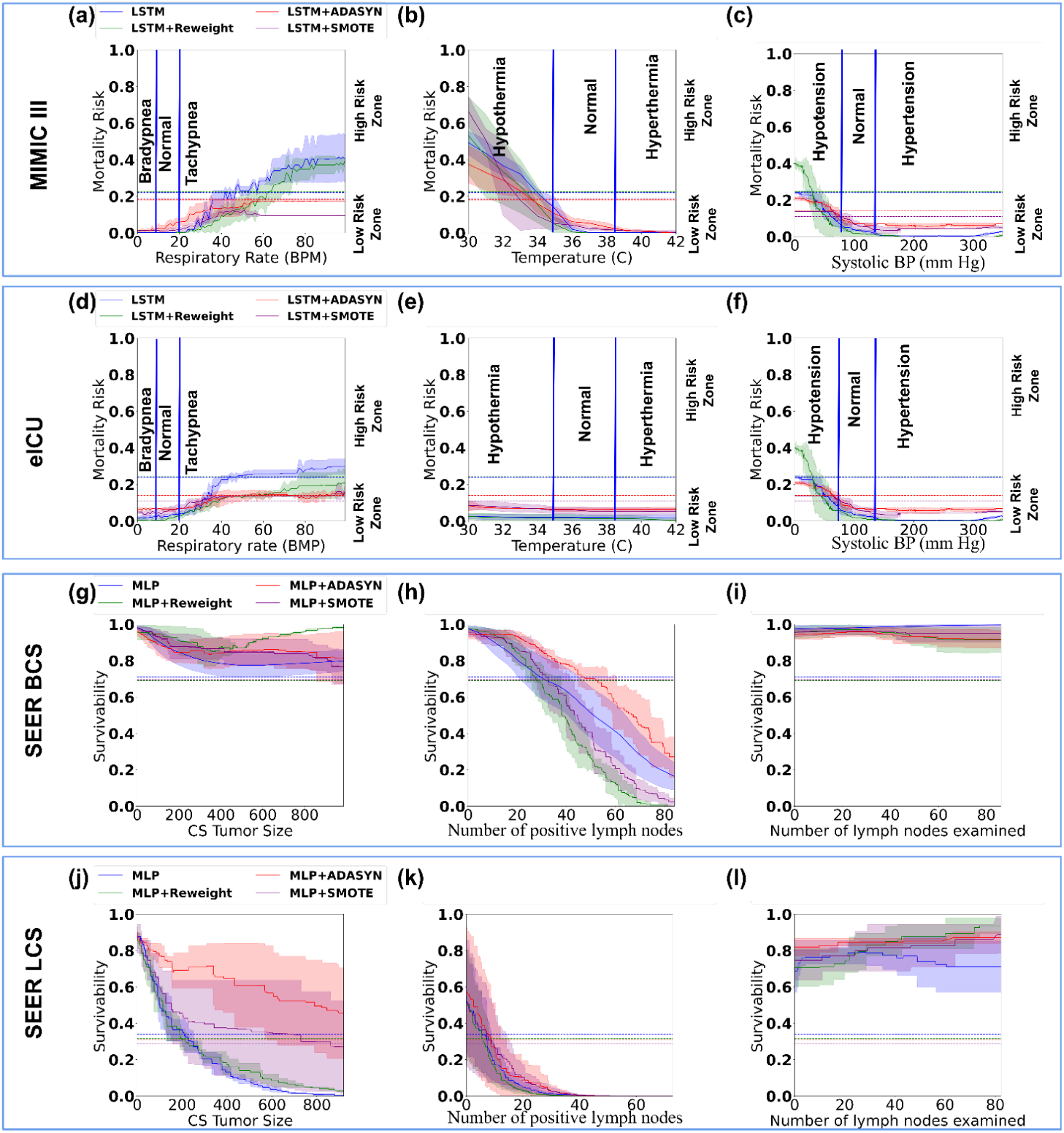
Performance comparison between the original machine learning models and the resampled (SMOTE or AdaSyn) or reweighted models under single-attribute varying tests. (a)-(c) and (d)-(f) Mortality risk prediction results by the original LSTM model and the resampled or reweighted LSTM models under MIMIC-III and eICU test cases for respiratory rate, temperature, and systolic blood pressure, respectively. (g)-(i) and (j)-(l) 5-year cancer survivability prediction results by the original MLP model and the resampled or reweighted MLP models under SEER BCS and LCS test cases for CS tumor size, the number of positive lymph nodes, and the number of lymph nodes examined, respectively. Horizontal dashed lines represent model-specific thresholds.

*Tree-based ensemble methods*, including AdaBoost, Random Forest, and XGBoost, produced somewhat similar results across all four datasets (Supplementary Figure S4). The random forest model demonstrates good responsiveness for most attributes in both MIMIC III and eICU datasets. However, it does not respond to critically high respiratory rates. XGBoost and AdaBoost have little reaction to attribute changes. In cancer survivability tasks, none of the models responds to worsening patient conditions, except AdaBoost for LCS-positive lymph nodes.

#### Double-attribute test results

The BCS-MLP model was tested under 60,462 double attribute varying test cases, including *i)* tumor size (T) and positive lymph node (N) combination test (Figure 6d), *ii)* number of examined lymph node (ENL) and positive lymph node (N) combination test (Figure 6e), and *iii)* tumor size (T) and number of examined lymph nodes (ENL) combination test (Figure 6f). The predicted survivability decreases with an increasing number of positive lymph nodes. However, collaborative staging (CS) tumor size or number of examined lymph nodes does not significantly decrease the predicted survivability. The MLP model accurately predicts 93% of T-N cases, 19.6% of N-ENL cases, and 0% of T-ENL cases (Supplementary Table 18). In a 3-attribute varying test, the BCS-MLP model is evaluated using cases with T4 tumor size, N3 number of positive lymph nodes, and grade 4 condition at the same time (Figure 6h). The BCS-MLP accurately predicts 90% of cases generated from surviving seeds (Class 1) and 98.9% of cases generated from death event seeds (Class 0).

#### Comparison of Wasserstein distances

We computed the Wasserstein distance between the original dataset and the generated test cases. Wasserstein distance captures the probability distribution shift given a metric space^25,26^. For in-hospital mortality prediction, the Wasserstein distance between the original MIMIC-III training set and the synthesized multi-attribute tests is 33.4. This value is much larger than the Wasserstein distance (12.4) between the training data and test data split within the original MIMIC-III. In comparison, for breast cancer survivability prediction, the distribution shift of the generated triple-attribute-based test cases from the original SEER dataset is smaller, with the Wasserstein distance being 9.8. WD for the original SEER training and test set is 2.1 (Supplementary Table 19).

### Impacts of resampling and reweighting methods

We trained and tested new ML models to assess the impact of resampling and reweighting methods on models’ responsiveness. SMOTE and AdaSyn oversampling methods are used to enrich the minority prediction class. For MIMIC-III mortality prediction, resampled LSTM models are tested with our single-attribute critical zone test cases. Overall, the new models remain to have low responsiveness to high-risk patient conditions (Figure 7a-7c). Similar to the original models, models with resampling are still unable to recognize critical patient conditions. For example, LSTM with SMOTE consistently assigns low mortality risk scores to patients with critically high vitals (e.g., respiratory rate, temperature, systolic blood pressure). LSTM with AdaSyn is better at responding to elevated systolic blood pressures than the original model, however, it performs poorly in other tests. Tests with the eICU dataset give a similar or worse performance (Figures 7d-7f). For BCS and LCS prediction, new MLP models trained with SMOTE or AdaSyn oversampling methods exhibit similar trends as the original MLP model (Figures 7g-7l). The new models fail to recognize many critical cancerous conditions. In addition, for LCS prediction, SMOTE and AdaSyn methods make the LCS-MLP model less sensitive to increasing CS tumor size (Figure 7j).

We also applied the reweighting approach to training. Supplementary Table 5 shows the cost parameters used. For mortality prediction, LSTM with reweighting gives comparable performance to the original model (Figures 7a-7f), except in one testing scenario. For eICU critically low systolic BP tests, reweighted LSTM generates elevated risk scores and is slightly better at responding to abnormal patient conditions. However, reweighted LSTM performs worse for similar MIMIC-III test cases (Figure 7c). For cancer survivability prediction, reweighting does not impact MLP’s performance in most testing scenarios (Figures 7g-7l). For BCS test cases, the reweighted MLP model has slightly better responses to the increasing number of positive lymph nodes than the original MLP, however, it performs worse than the original MLP in terms of recognizing larger tumor sizes.

### Responsiveness results of transformer models

The transformer models exhibit more responsiveness than LSTM in mortality prediction. They show elevated response in critically high zones for respiratory rate and systolic blood pressure, as well as in the critically low zone for temperature (Figure 8). This trend is observed for the single-attribute test cases of both MIMIC-III and eICU datasets. In addition, transformer models recognize both critical zones of systolic blood pressure, yielding a desired U-shaped response curve (Figures 8e and 8h). However, the transformer models fail to recognize critically low respiratory rates and critically high temperatures and exhibit low responsiveness to critically low systolic blood pressure. It also has delayed response to abnormally high respiratory rates. The transformer model’s risk prediction fluctuates significantly for eICU test cases.

**Figure 8:**
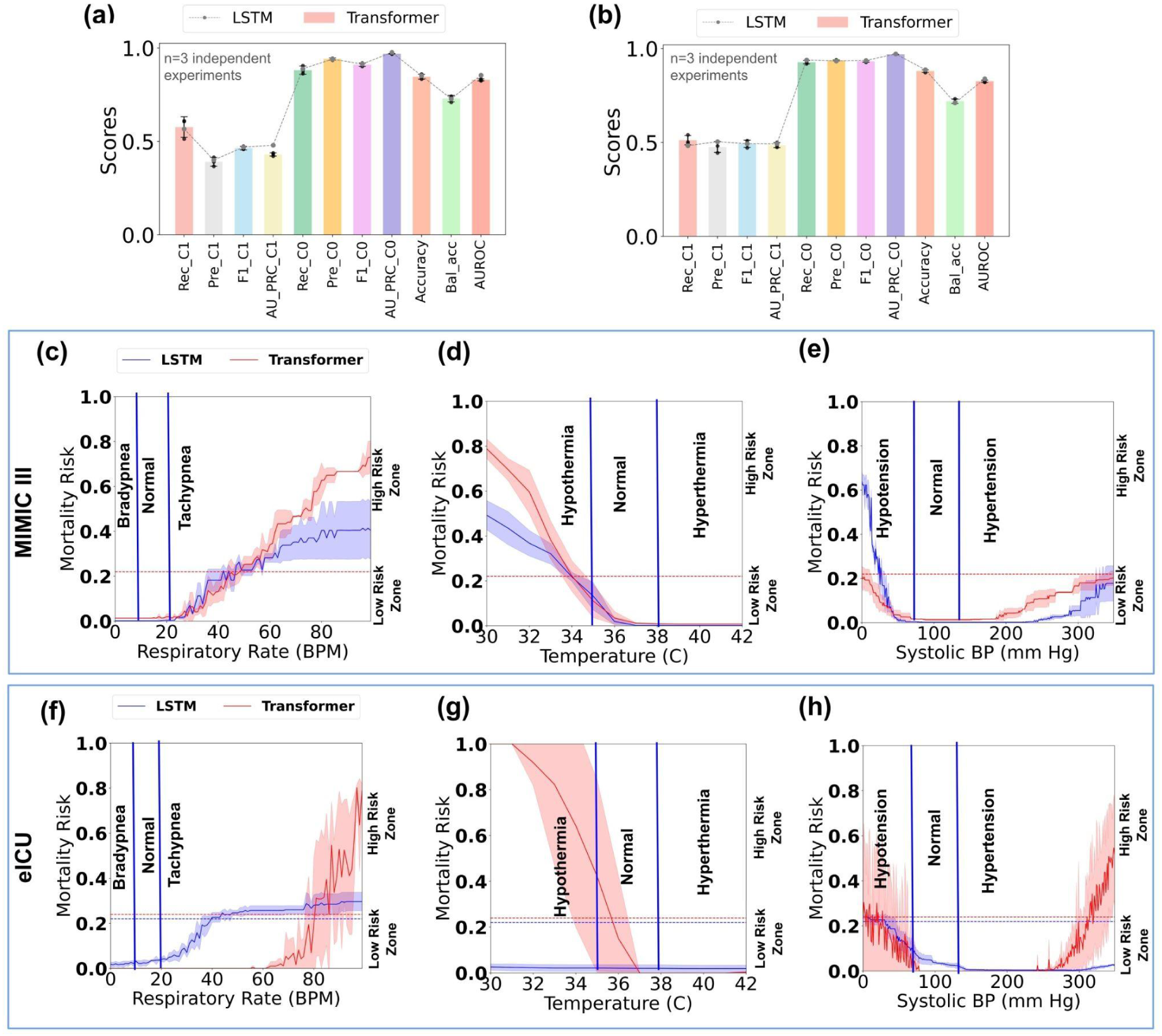
Performance and responsiveness of the transformer model compared with LSTM. Figures (a) and (b) show the various Class 1 (death) and Class 0 (survival) performance of the transformer model trained and tested on the original MIMIC-III and eICU datasets, respectively, with error bars indicating the standard deviation from three experimental trials. The dashed line represents the LSTM model’s performance. Figures (c)-(e) show the predicted mortality risk by the transformer model for respiratory rate, temperature, and systolic blood pressure on MIMIC-III single-attribute test cases, while (f)-(h) show the same for eICU test cases. Horizontal dashed lines denote model-specific thresholds for mortality risk prediction. Rec_C1, Pre_C1, F1_C1, AU_PRC_C1, Rec_C0, Pre_C0, F1_C0, AU_PRC_C0, Accuracy, Bal_Acc, and AUROC stand for Recall Class 1, Precision Class 1, F1 score Class 1, Area Under the Precision-Recall Curve Class 1, Recall Class 0, Precision Class 0, F1 score Class 0, Area Under the Precision-Recall Curve Class 0, Accuracy, Balanced Accuracy, and Area under the Receiver Operating Curve, respectively.

## Discussion

Our findings highlight the importance of measuring how clinical machine learning (ML) models respond to serious patient conditions. Our results show that most ML models tested are unable to adequately respond to patients who are seriously ill, even when multiple vital signs are extremely abnormal. For time-sensitive in-hospital mortality prediction, the lack of response to disease conditions is particularly troublesome. ML responsiveness is somewhat related to feature importance in some cases, e.g., the low responsiveness of LSTM to oxygen saturation tests (Figure 3f and Supplementary Table 16) is consistent with that feature’s low (15^th^) ranking (Supplementary Figure S5). However, for high-ranking features such as glucose and temperature, ML responsiveness to them is still inadequate. This poor responsiveness is also observed in the lack of responses in neural activation values (Figure 3 and Supplementary Table 17) to important vital changes, such as extremely low respiratory rate or high body temperature.

New ML responsiveness metrics, especially for the healthcare domain, are urgently needed. ML responsiveness is a new problem. It differs from the well-studied ML robustness^40^. ML robustness aims to ensure model stability and the ability to resist sample perturbations so that small (maliciously injected) noises to samples cannot change the prediction results. Lipschitzness, a common ML robustness metric, measures the model’s resilience to noisy data and perturbations^41^. However, for healthcare applications, optimizing Lipschitzness may lead to models being even more insensitive to changes in patient conditions, as adherence to Lipschitz continuity may hinder the model’s ability to capture crucial input variations. In image and natural language domains, a common testing approach is adversarial attacks^42–45^. That testing approach involves intentionally manipulating input data to deceive the model’s predictions and does not apply to our medical settings.

Our results identified serious deficiencies in conventionally trained binary classification models in recognizing seriously abnormal medical conditions. For example, in-hospital mortality prediction models fail to generate alerts for bradypnea (low respiratory rates) or hypoglycemia conditions (Figure 3). Similarly, the models also consistently underestimate some of the mortality risks when given multiple abnormal vital time series in conjunction (Figure 4). When given test cases representing various injury levels, neural network models (namely, LSTM and CW-LSTM) gave inconsistent risk predictions -- assigning higher mortality risk (> 0.5) to cases of moderate injury (e.g., GCS score 12), while assigning disproportionately lower risk (< 0.05) to severe injuries (e.g., GCS score 7). The two neural network models exhibit insensitivity to changes in eye response (Figures 2d and 2f). For most attributes, we found the training data’s distribution is highly centered, not sufficiently representing high or low critical zones (Supplementary Figure S1). Death and non-death cases exhibit somewhat similar value distributions, means, and standard deviations (Supplementary Table 14) for individual attributes, despite the drastically different outcome. Machine learning methods produced by supervised training approaches are unable to recognize the meanings of vitals in dangerous zones. This semantic deficiency of ML models was also reported in image recognition studies, e.g., melanoma classification overinterpreting surgical skin markings^8^. We found similar kinds of semantic deficiencies in an ML model predicting 5-year breast cancer survivability. These findings indicate the importance of empirically assessing the trustworthiness of clinical ML models.

The conventional test set is limited in its distribution shift from the training data. For example, for in-hospital mortality prediction, our generated multi-attribute test cases present a high Wasserstein distance (33.4) from the original MIMIC-III training data, much larger than the split test set’s Wasserstein distance (12.4). A similar distribution shift pattern is observed for the breast cancer survivability model. For triple-attribute test cases, the breast cancer survivability prediction model performs better (89-98% triple-attribute test accuracy) than in-hospital mortality prediction models (6-69% multi-attribute test accuracy). This difference in accuracy may be partly due to the different distribution shift in generated test data. There is a much smaller distribution shift in triple-attribute breast cancer test cases (Wasserstein distance 9.8) than in multi-vital test cases (Wasserstein distance 33.4), with respect to their original training data. The LSTM model is slightly better at recognizing multi-attribute test cases (45.7% accuracy) than single-attribute ones (37.8%) and CW-LSTM exhibits a similar pattern (69.3% vs. 22.4%, Supplementary Table 16). Multiple abnormal vitals likely provide more clues for the ML models to classify, whereas single isolated attribute changes appear more difficult. The poor performance of the ML models is somewhat expected because of the distribution shift between training data and our synthetic test data. Yet, these deficiencies are unacceptable from a clinical deployment perspective, as the test cases represent potential real-life medical conditions. Our work points out a fundamental limitation of pure data-driven machine-learning models, which is *models purely trained by patient data do not perform well for tasks that require implicit medical knowledge (e.g., normal vital ranges)*.

For in-hospital mortality prediction, all models have multiple deficiencies under single-attribute critical zone test cases and are unable to generate high enough risk predictions for serious patient conditions (Figures 3, 8, and Supplementary Table 16). Out of all the dual critical zone attributes, only LSTM and transformer models exhibit U-shape risk curves for systolic blood pressure (Figures 3e and 8e). The other risk curves are either monotonic or flat (Figure 3). Transformer models implementing the parallel attention mechanism are known to be better at capturing the global context and dependencies in data than sequential models like LSTM. Indeed, transformers are more responsive to abnormal vitals than LSTM in single-attribute testing (Figure 8). However, our results suggest advanced models alone are not sufficient, as there are still multiple unrecognized critical zones. For multiple attribute testing, logistic regression performs the worst (Figure 4 and Supplementary Table 16), mispredicting 93.7% of test cases.

CW-LSTM’s accuracy is the lowest (22.4%) in single-attribute testing, however, it gives the highest average accuracy (69.3%) for multiple-attribute testing. Brain injury-related Glasgow Coma Scale test cases (Figure 2) involve simple categorical data (as opposed to numerical data). Logistic regression gives the best performance, generating appropriate and consistent risk estimates, and substantially outperforms the two neural network models (Supplementary Table 16). These results suggest that for categorical attributes such as GCS, a simpler model like logistic regression may be more suitable than complex deep learning models, indicating the importance of evaluating a wide variety of machine learning models before clinical use. Deficiencies in ML responsiveness were also observed in the 5-year breast cancer prediction task -- the MLP model gave an average of 48.9% prediction accuracy under our test case (Supplementary Table 18). This accuracy is much lower than the widely reported death class accuracy of 90% (standard deviation 0.45), computed based on the original test data from SEER^5,19^.

The linear logistic regression model tested is unsuitable for analyzing vitals due to multiple reasons. It reduces time series to statistical summaries (Supplementary Figure S6c) and is unable to capture data dynamics. Linear logistic regression is unable to model non-linear (e.g., U-shaped curve) features, as it responds monotonically to features. In multi-attribute tests, the model gives poor performance (6.2% accuracy, Supplementary Table 16), partly because of its many negative coefficients associated with attributes. 13 out of the 17 attributes have negative coefficients, e.g., the temperature is strongly inversely correlated with the predicted probability (Supplementary Figure S6a), resulting in underestimated risk prediction. Gaussian Naive Bayes and KNN show weaker performance on the original test sets than the others (Supplementary Figure S7) and, thus, are excluded from subsequent attribute tests. For completeness, model performance on the original test set was given in Supplementary Figure S7.

For in-hospital mortality prediction, the LSTM models are slightly better at recognizing deteriorating trends (average 44%, Figure 5) than cases with steadily low or steadily high vitals in critical zones (average 36.5%, Figures 3 and 4). When test cases contain 3 simultaneously deteriorating attributes, the models detected 56% of them on average, which is better than their performance of 41% on a single deteriorating attribute. When using LSTM gradient ascent to automatically generate multi-attribute deteriorating test cases, we found the resulting test cases all have significantly decreased oxygen saturation and body temperature values in the last 24 hours. Because the gradient ascent process follows the shortest path within the loss function space of the model, these findings indicate that *i)* oxygen saturation and body temperature are top LSTM features and *ii)* the last 24 hours (out of the entire 48-hour timespan) are important in the model’s decision-making process, which is also consistent with logistic regression feature ranking (Supplementary Figure S6b).

The BCS multilayer perceptron model (MLP) model exhibited responsiveness to critical attributes, such as tumor size and lymph node involvement (Figure 6). For example, for N3 stage (extensive lymph node involvement) test cases, the model was able to raise alerts for 74.4% of them (Supplementary Table 18). The model also performed well (nearly 100% alerts) when all three critical features (T4 tumor size, N3 lymph node stage, and grade 4) were high. These observations suggest the MLP model’s prediction capability in extreme cases is good. However, the model does not respond to severe tumor sizes (T3 stage), generating no alerts. The consistency in predicted survivability scores is also low, as the model generated slightly more alerts for grade 3 cancer (65.8%) than grade 4 terminal cancer (63.9%). This inconsistency may be the outcome of the imbalanced dataset (Supplementary Figure S2), as the SEER dataset contains a total of 81,749 (death 15,628 and survived 66,121) grade 3 cases, while only 3,002 (death 640 and survived 2,362) grade 4 cases. The number of examined lymph nodes (ELNs) does not directly indicate a cancerous condition, thus, the model’s lack of response to ELN is somewhat expected.

Despite overall good performance on the *original* test set (Supplementary Figure S7), tree-based ensemble methods such as XGBoost, AdaBoost, and Random Forest exhibit low responsiveness to critical zone tests (Supplementary Figure S4). Ensemble methods perform much worse than MLP for SEER BCS and LCS settings, which is likely due to the sparsity in the one-hot encoded input space. The SEER dataset has much larger feature dimensions (56 for BCS and 47 for LCS) than the MIMIC III and eICU time-series data (17 features). Using the one-hot encoding to encode categorical features leads to an expansive number of sparse encoded representations (1,423 for encoded BCS and 1,315 for encoded LCS), posing challenges to tree-based models.

One clinical mitigation is to deploy a filter-then-predict workflow where domain-specific rules are first applied to identify cases with obvious disease conditions. Thus, corner-case scenarios will never reach machine learning models. However, designing such rule-based classifiers, especially under time-series data, is challenging and may require substantial manual efforts. A more efficient approach is out-of-distribution detection, which identifies cases that present a significantly large distribution shift from the model’s training data. Existing solutions for detecting out-of-distribution images^46^ cannot be directly applied to clinical settings. For medical applications, out-of-distribution patient cases still need to be examined and handled. An overly strict detection may produce too many such out-of-distribution cases for downstream examination. Finding the right balance will facilitate clinical translation.

A promising direction is medical foundation models based on clinical large language models (LLMs)^47–50^. Our findings suggest that statistical machine-learning models solely trained from patient data are grossly inadequate. They are unable to capture basic clinical knowledge, e.g., patients with extremely low Glasgow Coma Scale values have a high mortality risk. LLMs are likely able to recognize common sense health conditions and serve as a filter mechanism before ML classifiers. However, it is crucial to quantitatively characterize the trustworthiness of medical LLMs before clinical adoption. Our work suggests the urgent need for innovative clinical decision-making workflows, as existing models solely trained from patient samples are extremely limited. At clinical time, a human-friendly interface is also important for interpreting ML results. Conventional interpretability techniques, such as SHapley Additive exPlanations (SHAP)^51^, Local Interpretable Model-Agnostic Explanations (LIME)^52^, or TRUSTEE^53^, were designed for ML experts, not for clinicians. Therefore, these tools cannot be directly used in clinical settings. An innovative clinical workflow needs to place large language models (LLMs) as the final component to generate narrative explanations based on ML predictions and interpretability results. An interesting research direction is how to fine-tune LLMs for these specific tasks.

The boost provided by conventional resampling and reweighting methods is very limited (Figure 7). Under some scenarios (e.g., tachypnea and increasing CS tumor size), they may perform even worse than the original models. This poor performance is expected, as these methods rely on existing minority class samples in the training set, which are limited in their ranges and variations. The root problem is that the space of all possible minority class samples is vast. Attempting to cover all or most of them through training data engineering (such as oversampling) is infeasible. Thus, data engineering does not appear to be a feasible direction for the ML responsiveness problem. A more promising approach is to directly encode medical semantics into the clinical decision workflow as discussed above.

Our work provides the first look into ML responsiveness. Comprehensive measurement studies in other medical settings are needed. Our gradient ascent testing methodology can be extended to other health conditions (e.g., rare diseases or comorbidities). Scalability is the key to testing in medicine, because of the complex high-dimensional space. Innovative methods that prioritize testing are needed to reveal the most critical blind spots in a model.

## Data availability

The MIMIC-III, eICU, and SEER data used in this study are not publicly downloadable, but can be requested at their original sites after completing proper training. Parties interested in data access should visit the MIMIC-III website (https://mimic.physionet.org/gettingstarted/access/), eICU website (https://eicu-crd.mit.edu/) and the SEER website (https://seer.cancer.gov/data/access.html) to submit access requests. Because our test cases are generated from these access-controlled datasets, they cannot be publicly released. However, we have released the code for reproducing all our test cases. All the source-data for the main figures (as a Microsoft Excel file) is available as Supplementary Data 1.

## Code availability

We have released all our code publicly on GitHub, which can be used to generate the test cases and reproduce our experiments. https://github.com/PiasTanmoy/TRUSTWORTHY-ML (https://doi.org/10.5281/zenodo.14254248)^54^

## Contributions

D.Y. conceived and designed the study. T.P. and D.Y. conceived the gradient-based test generation method. D.Y. and T.P. conceived the new metrics introduced. T.P. conducted the experiments and analyzed the data, including selecting seeds for test case generation and interviewing medical experts. S.A. performed the initial neuron heat map study. X.D. guided T.P. to perform the WD statistical analysis. M.T. and I.T. provided medical feedback on the mortality risks of generated test cases through Zoom interviews. D.Y. and T.P. wrote the manuscript. C.B.N. provided strategic guidance. All authors proofread the manuscript and provided feedback.

## Supplementary Materials

**Supplementary Figure S1.**
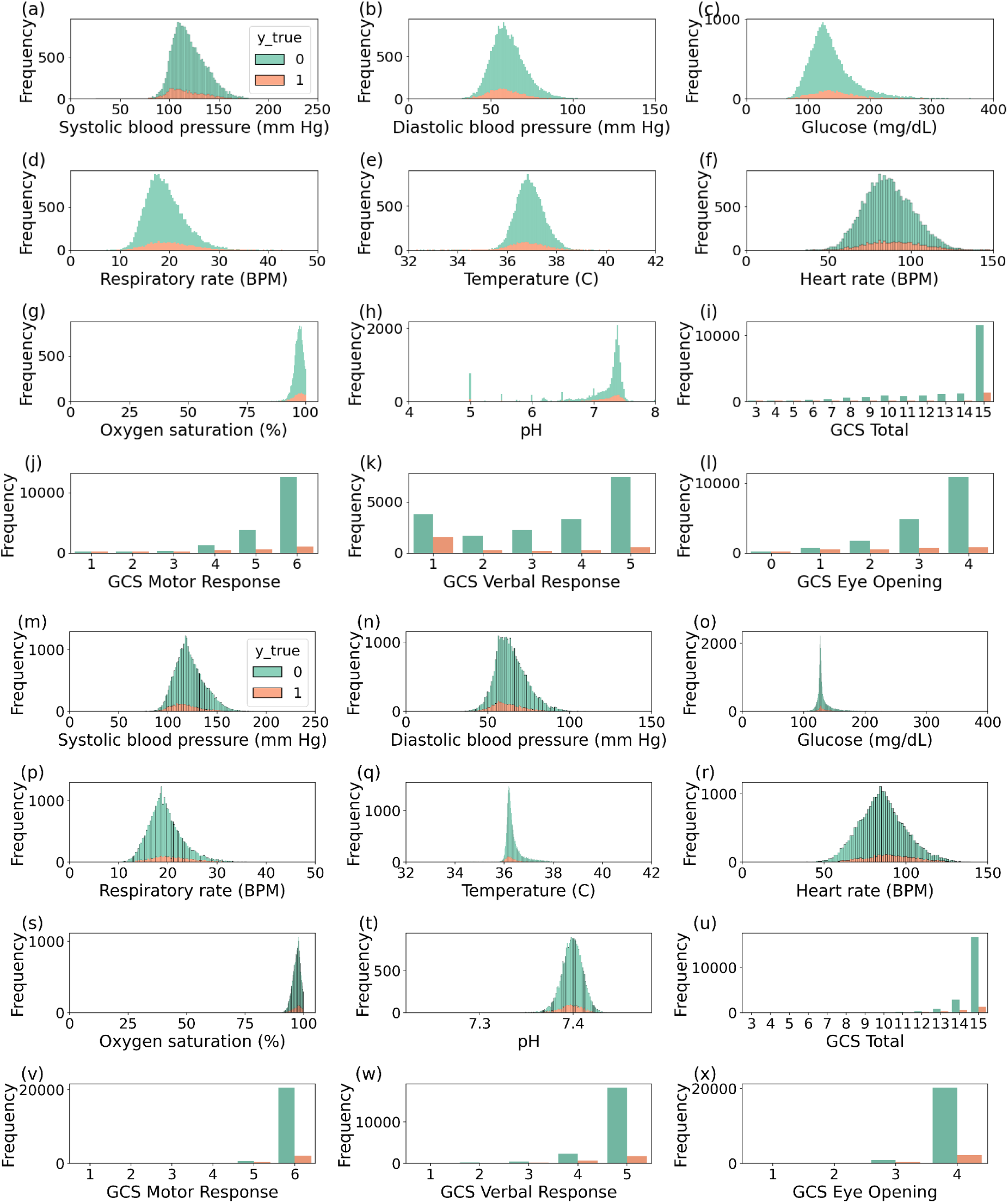
Distributions of different attributes (vitals) of the (a)-(l) MIMIC-III and (m)-(x) eICU 48 hours ICU mortality dataset.

**Supplementary Figure S2:**
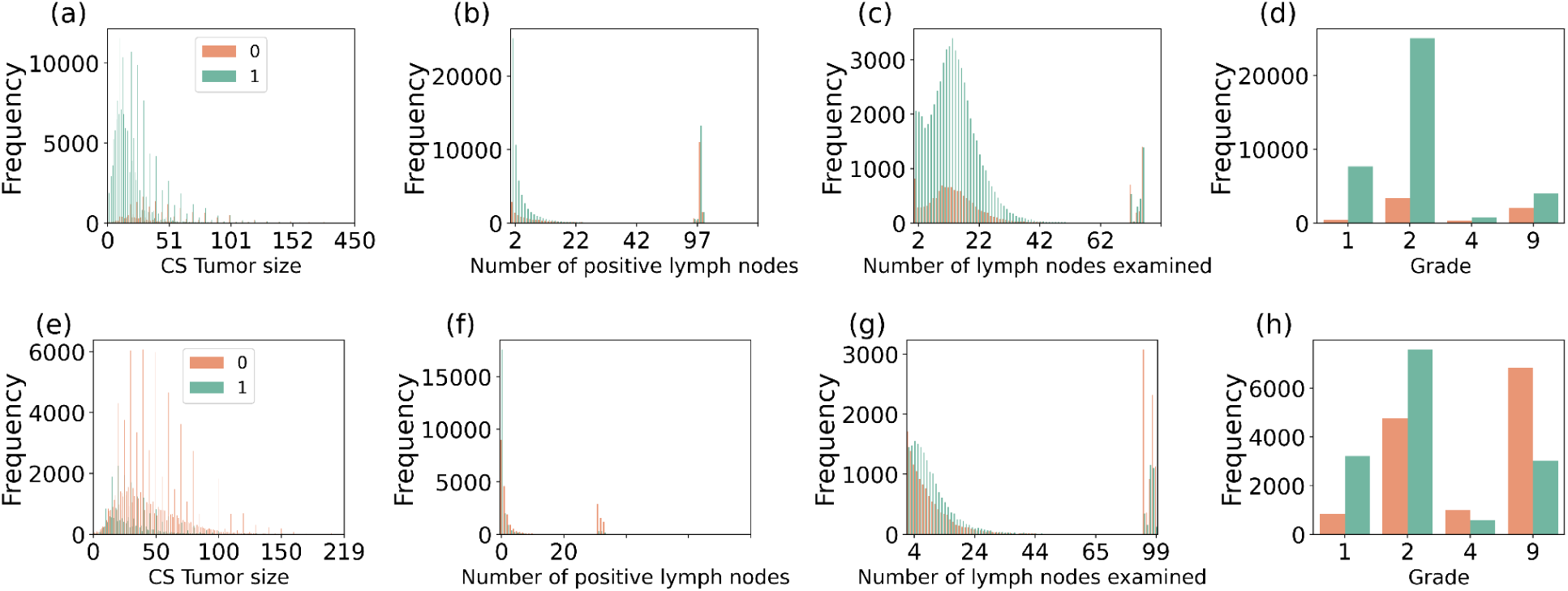
Distributions of the SEER 5-year (a)-(d) breast and (e)-(h) lung cancer survivability dataset. Here label 0 represents the death case and 1 is mapped to the survival case. Death cases are the majority in the LCS dataset, whereas survival cases are the majority in the BCS dataset.

**Supplementary Figure S3.**
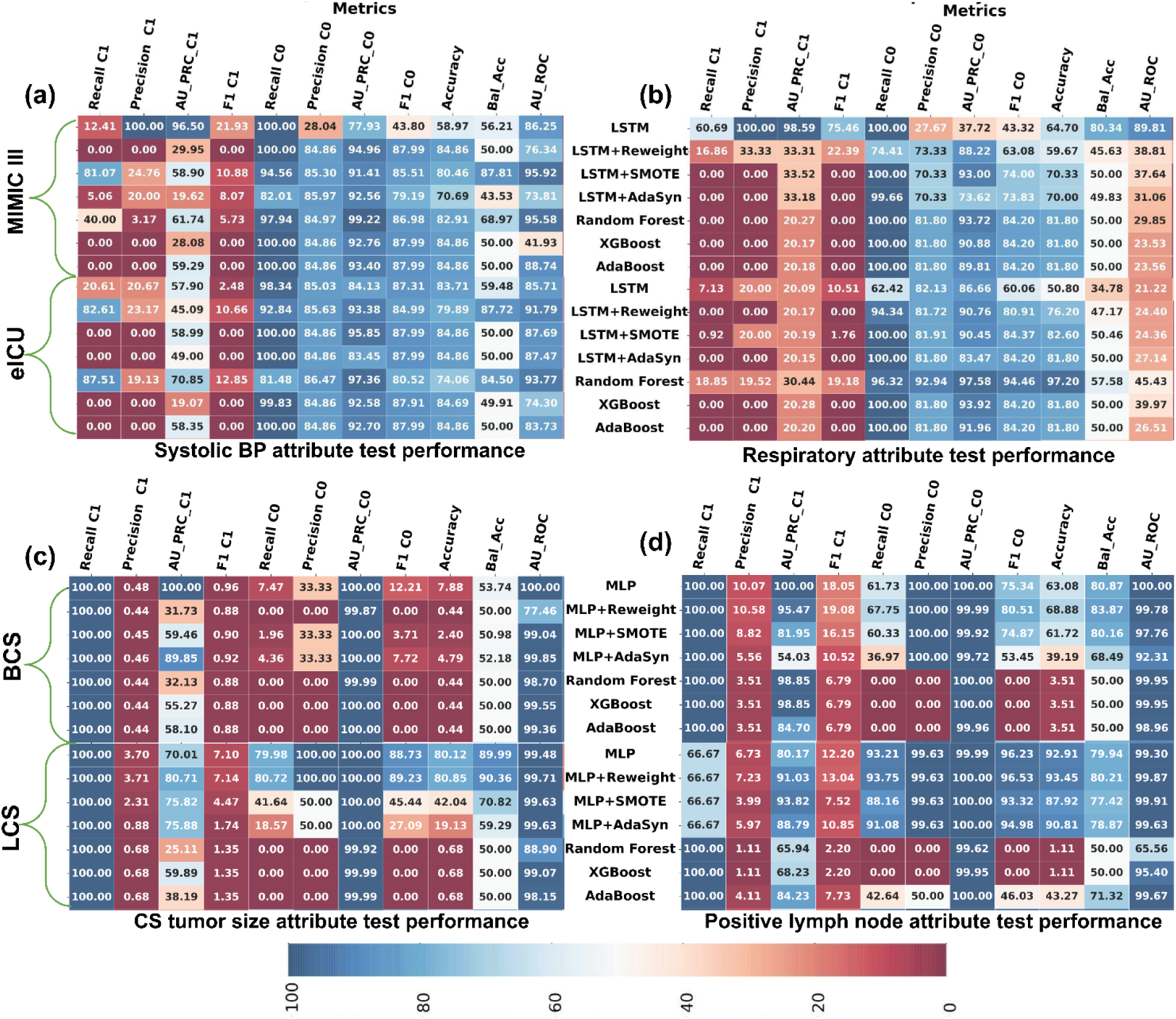
C0 and C1 represent class 0 and class 1 respectively. AU_PRC, Bal_Acc, and AU_ROC represent area under Precision-Pecall Curve, Balanced Accuracy, and Area Under Receiver Operating Curve, respectively. The performance of machine learning models was evaluated on synthesized single-attribute test sets, including (a) ICU vital systolic blood pressure, (b) ICU vital respiratory rate, (c) cancer attribute CS tumor size, and (d) cancer attribute number of positive lymph nodes. Models were trained on the original datasets, with the top halves of (a) and (b) representing MIMIC III and the bottom halves representing eICU. For (c) and (d), the top halves represent SEER BCS and the bottom halves represent SEER LCS, as indicated on the left. Model names are aligned along the Y-axis. For ICU mortality prediction (a) and (b), Class 1 (C1) corresponds to the death class, whereas for SEER cancer survivability prediction (c) and (d), Class 0 (C0) represents the death class.

**Supplementary Figure S4:**
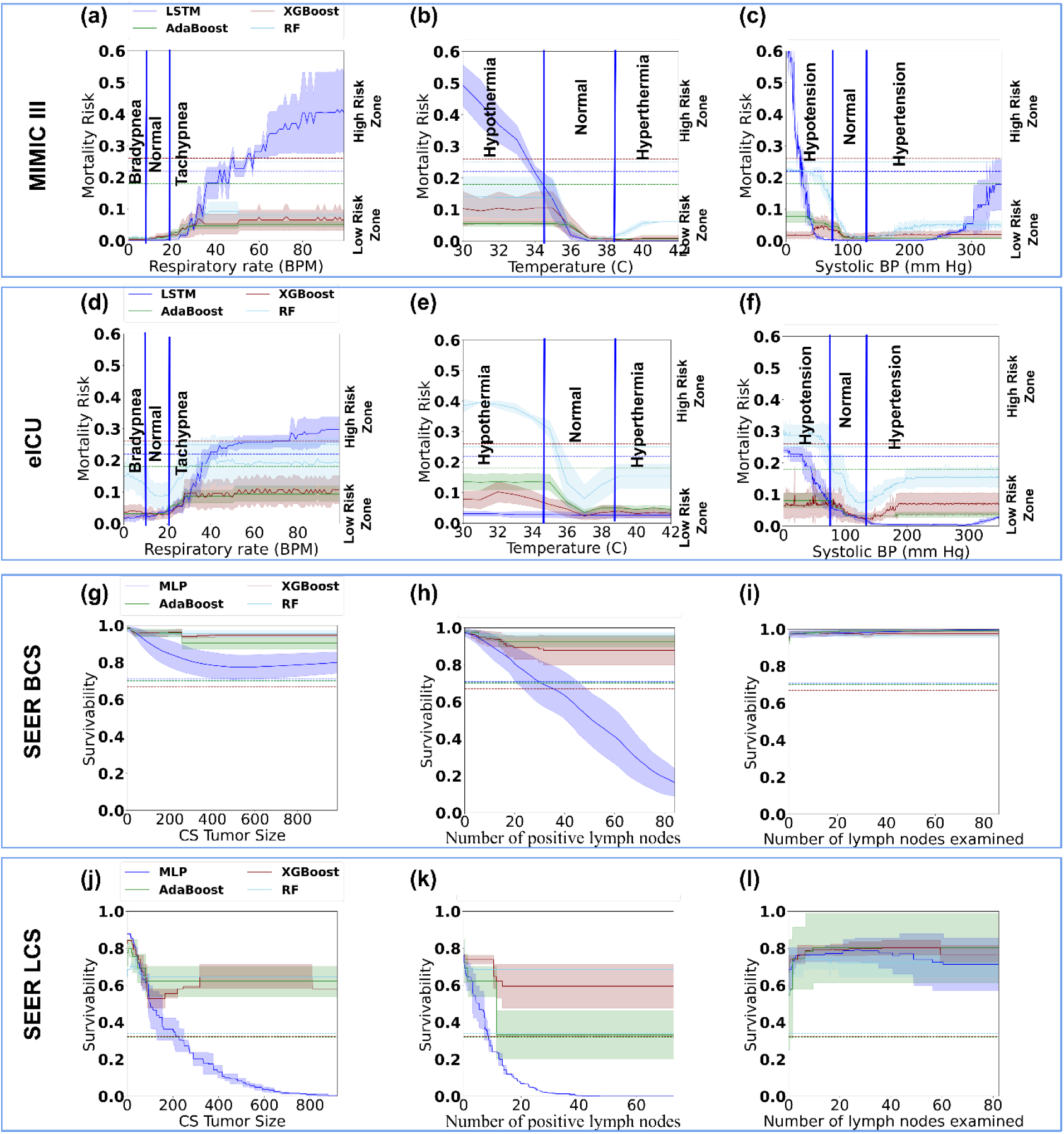
Performance comparison between tree-based ensemble methods, AdaBoost, XGBoost, and Random Forest (RF), with LSTM and MLP models under single-attribute varying tests. (a)-(c) and (d)-(f) Mortality risk prediction results by the models under MIMIC-III and eICU test cases for respiratory rate, temperature, and systolic blood pressure, respectively. (g)-(i) and (j)-(l) 5-year cancer survivability prediction results by the models under SEER BCS and LCS test cases for CS tumor size, the number of positive lymph nodes, and the number of lymph nodes examined, respectively. Horizontal dashed lines represent model-specific thresholds.

**Supplementary Figure S5:**
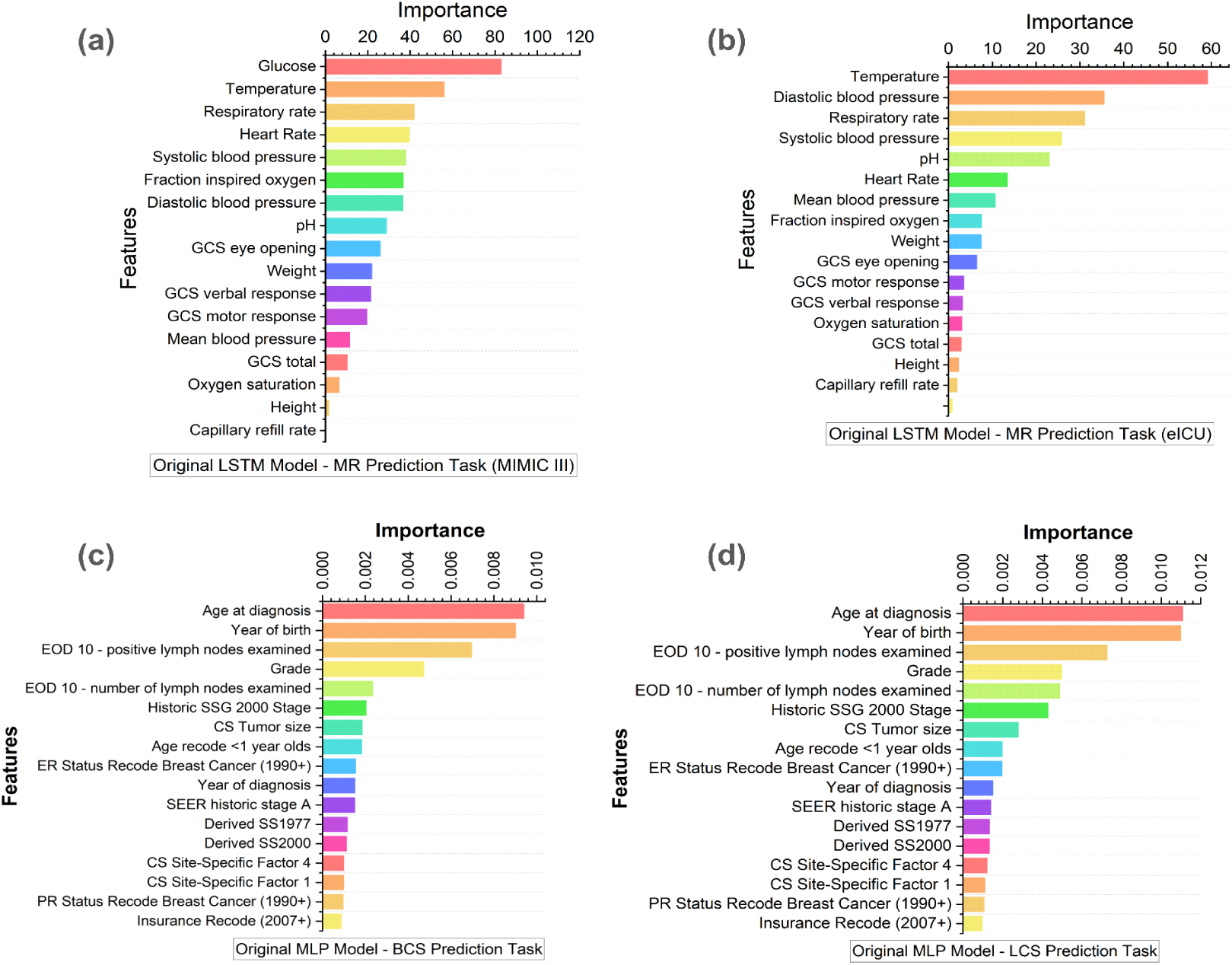
SHAP-average (of one-hot encoded features) feature importance of the (a) LSTM trained on MIMIC III dataset, (b) LSTM trained on eICU dataset, (c) MLP trained on BCS dataset. (d) MLP trained on LCS dataset.

**Supplementary Figure S6:**
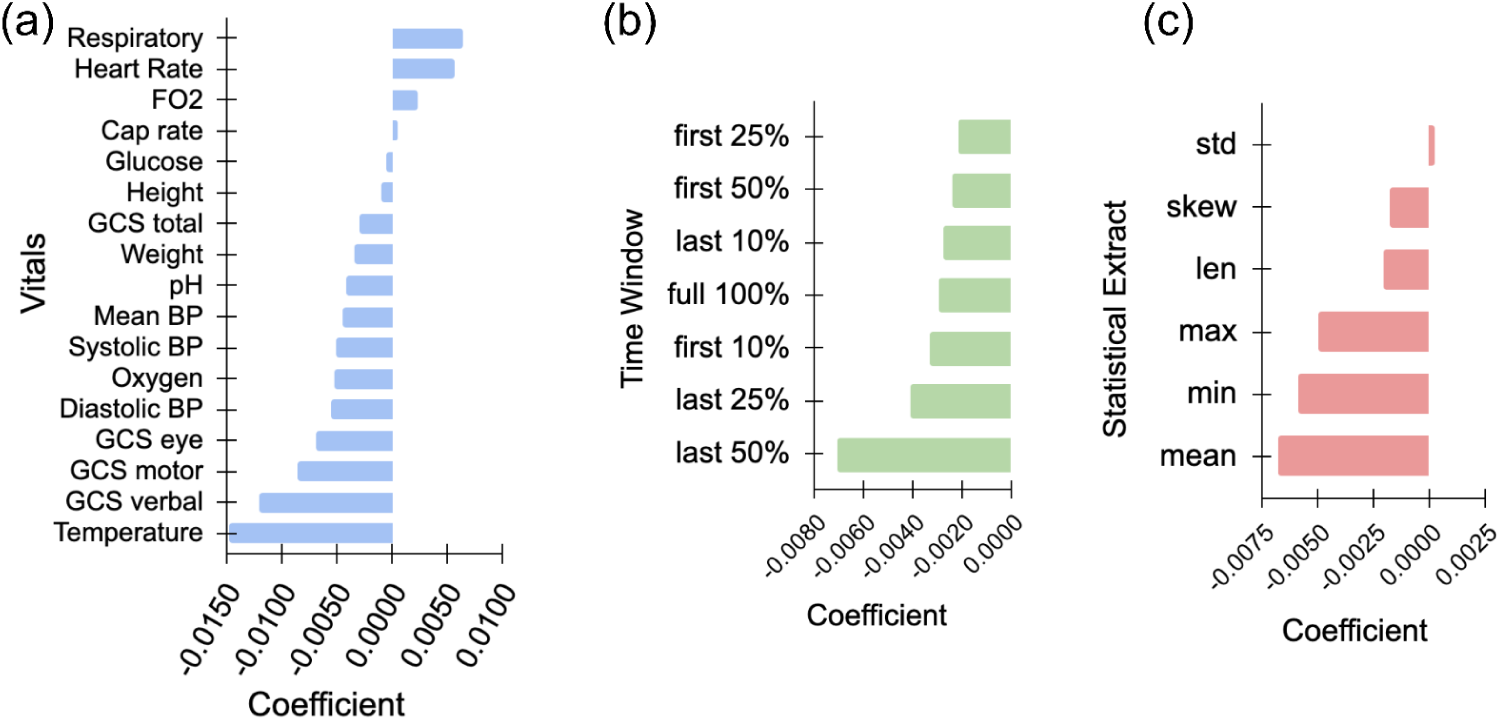
Logistic regression coefficients (averaged) for (a) vitals, (b) time-period, and (c) statistical-extract features.

**Supplementary Figure S7:**
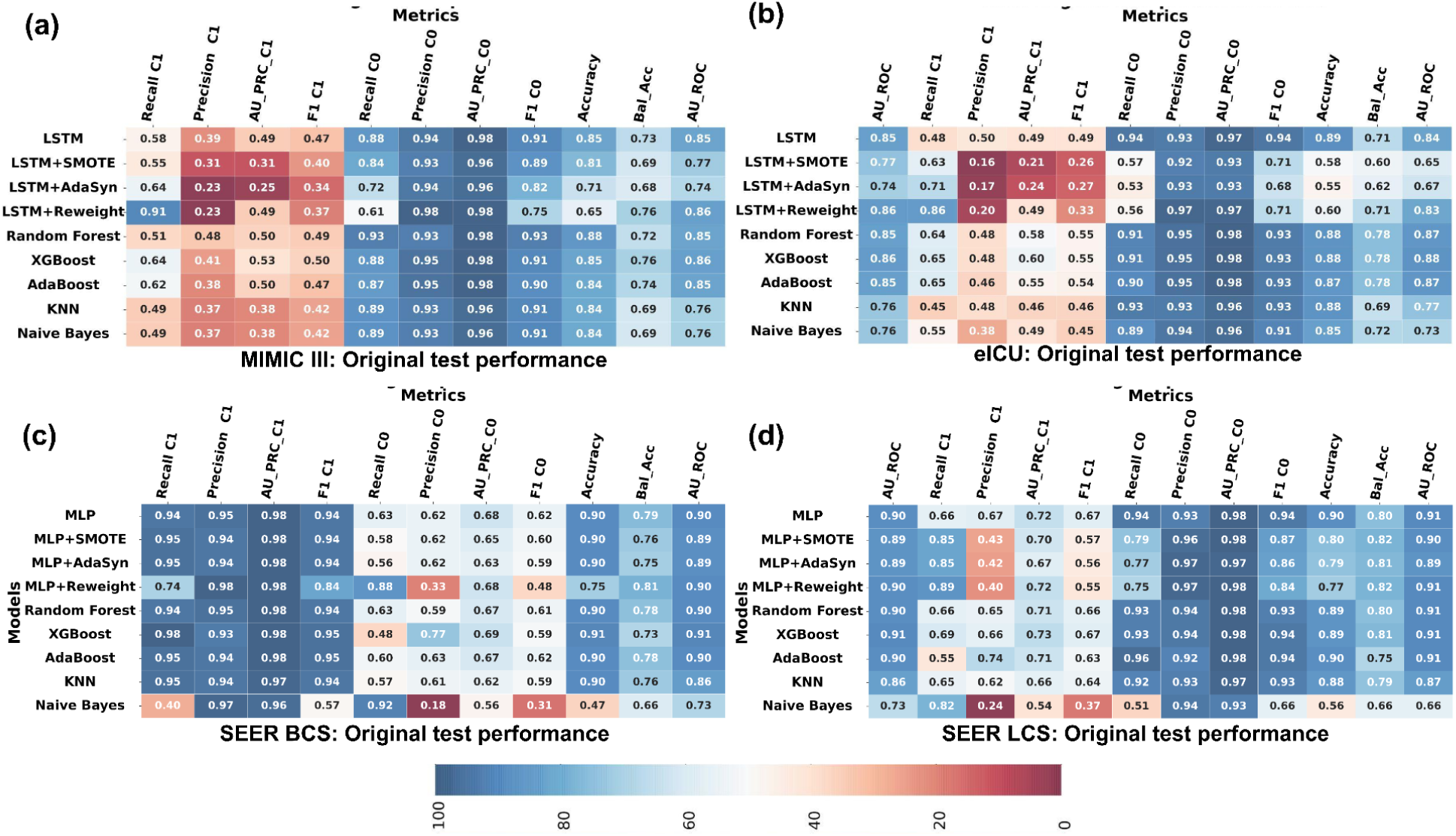
C0 and C1 represent class 0 and class 1 respectively. AU_PRC, Bal_Acc, and AU_ROC represent area under Precision-Pecall Curve, Balanced Accuracy, and Area Under Receiver Operating Curve, respectively. Machine learning model performance on the original test set. The death class is the minority class in MIMIC III, eICU, and SEER BCS datasets, whereas the majority in the SEER LCS dataset. Death class is represented by 1 in in-hospital mortality risk prediction datasets (MIMIC III and eICU) and 0 in cancer survivability prediction datasets (SEER BCS and LCS).

## Supplementary Notes

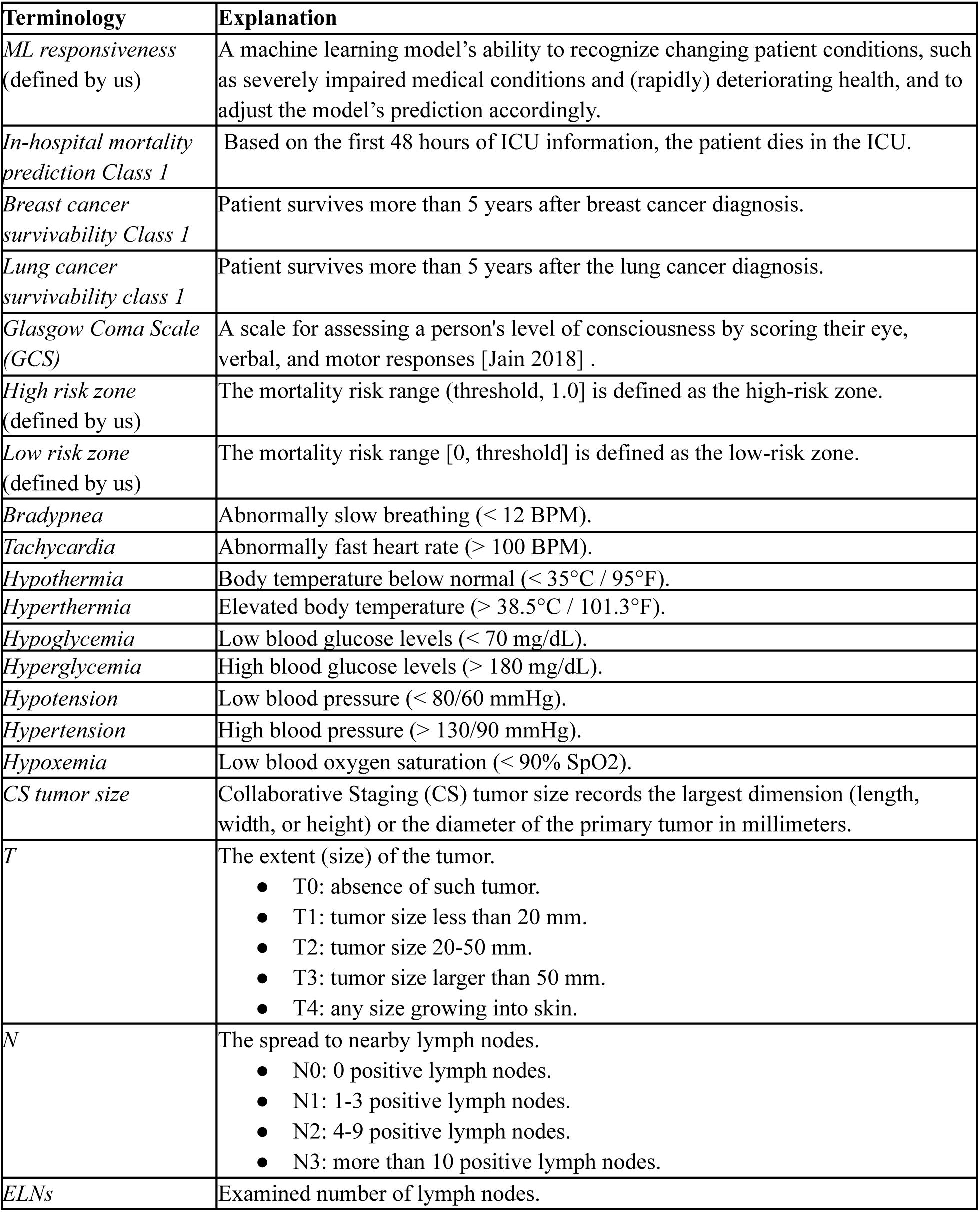

## Supplementary Equations

Given neuron *i*, its activation value *act*(*neuron*_i_ (*k*)) for input *k*, and a vital zone bounded by *n*_1_ and *n*_2_ (representing critically low, critically high, or normal vital range), we define the average neural zone activation *NZA_i_* (*n*_1_, *n*_2_) in Equation (1). Given two vital zones *z*_1_ and *z*_2_, we further define the average neural zone activation difference Δ*NZA*_avg_ (*z*_1_, *z*_2_) in Equation (2). In Equation (2), *z*_is_ and *z*_ie_ are the zone starting and ending values, respectively. *b* is the number of neurons in the activation layer (16 in our case).

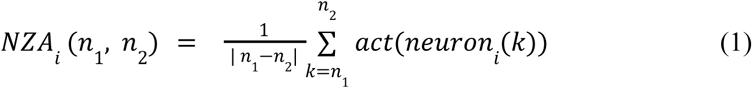

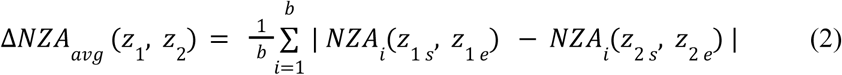

### Gradient ascent

Given a sample *x*, we add the gradient *G_i_* calculated from the trained model with the seed sample *x_i_* to generate a new sample *x_i_*^*new*^, where *i* represents the feature to be changed by the gradient ascent process.

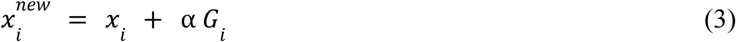

Where:

*i* is the selected feature index from [0, 1, …, 17]

*x_i_*^*new*^ is the sample with updated feature i

*x_i_* is the current feature value

α is the step size or learning rate that controls the magnitude of change in the input feature

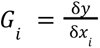 is the gradient of the output y with respect to feature *x*

## Supplementary Method

### Algorithm for Single-Attribute Variation Test Case Generation for Time Series Data

Input:

- Dataset D (e.g., MIMIC-III or eICU)
- Attribute A∈[*Diastolic bp, Glucose, Respiratory rate*, ....]
- Time-series length T = 48 hours
- Seed S

Output: Set of test cases with varied single attribute A Procedure:

1. Compute minimum *A_min_*, maximum *A_max_*, mean µ*_A_*, and standard deviation σ*_A_* values of attribute A in dataset D.
2. For each value μ in the range [*A_min_*, *A_max_*] with step size α

1. *x_A_* : Generate a vector of length T by sampling random values from the normal distribution 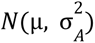
2. *S_µ_* : Replace the feature value of A with the new vector *x_A_* in seed S
3. Save the new sample, *S*_µ_ which is a representative of attribute value μ

---

**Supplementary Table 1:** Best hyperparameters of machine learning models selected using a grid search.

| Model | Hyperparameter search set | Best hyperparameters |  |  |  |
| --- | --- | --- | --- | --- | --- |
|  |  | *eICU* | *MIMIC-III* | *SEER-BC*<br>*S* | *SEER-LCS* |
| **XGBoost** | n_estimators: [50, 100, 150, 200, 300] | 200 | 300 | 300 | 300 |
|  | max_depth: [None, 3, 5, 7, 10] | 5 | 3 | 5 | 5 |
|  | learning_rate: [0.01, 0.1, 0.2, 0.5, 1] | 0.1 | 0.1 | 0.1 | 0.1 |
|  | subsample: [0.1, 0.5, 0.8, 1.0] | 1 | 1 | 0.8 | 0.8 |
|  | colsample_bytree: [0.1, 0.5, 0.8, 1.0] | 1 | 1 | 0.8 | 1 |
| **AdaBoost** | n_estimators: [50, 100, 150, 200, 300] | 200 | 200 | 150 | 150 |
|  | learning_rate: [0.01, 0.1, 0.2, 0.5, 1] | 0.1 | 0.1 | 1 | 1 |
| **Random Forest** | n_estimators: [50, 100, 150, 200, 300] | 300 | 300 | 300 | 300 |
|  | max_depth: [None, 3, 5, 7, 10] | None | None | None | None |
|  | min_samples_split: [2, 5, 10] | 10 | 10 | 2 | 10 |
|  | min_samples_leaf: [1, 2, 4] | 4 | 4 | 4 | 2 |
| **K Nearest Neighbors** | n_neighbors: [3, 5, 7, 9, 11] | 11 | 11 | 11 | 11 |
|  | weights: ['uniform', 'distance'] | distance | distance | distance | distance |
|  | metric: ['euclidean', 'manhattan', 'minkowski'] | euclidean | manhattan | euclidean | euclidean |
|  | p: [1, 2] | 1 | 1 | 1 | 1 |
| **Naive Bayes** | var_smoothing: [1e-9, 1e-8, 1e-7, 1e-6] | 1e-7 | 1e-6 | 1e-9 | 1e-9 |
| **Transformer** | number of heads: [2, 4, 8] | 4 | 4 | -- | -- |
|  | number of transformer blocks: [1, 2, 3] | 2 | 3 | -- | -- |
|  | dense units (transformer block): [16, 64, 256] | 16 | 16 | -- | -- |
|  | dropout: [0.1, 0.2, 0.3, 0.4, 0.5] | 0.5 | 0.3 | -- | -- |
|  | batch normalization: [True, False] | True | True | -- | -- |
|  | dense units: [16, 32, 64] | 32 | 64 | -- | -- |

**Supplementary Table 2:** Training and validation losses of selected in-hospital mortality risk predictor models.

| Prediction task | Model | Trial | Selected Epoch (1-100) | Validation AUPRC (C1) | Training Loss | Validation Loss | Difference (Training loss ~ Validation loss) |
| --- | --- | --- | --- | --- | --- | --- | --- |
| MIMIC III | *LSTM* | 1 | 39 | 0.568 | 0.273 | 0.278 | 0.005 |
|  |  | 2 | 29 | 0.564 | 0.279 | 0.278 | 0.001 |
|  |  | 3 | 33 | 0.566 | 0.274 | 0.279 | 0.005 |
|  | *CW-LSTM* | 1 | 37 | 0.556 | 0.270 | 0.284 | 0.014 |
|  |  | 2 | 24 | 0.556 | 0.280 | 0.286 | 0.006 |
|  |  | 3 | 20 | 0.561 | 0.283 | 0.281 | 0.002 |
|  | *LSTM+ SMOTE* | 1 | 4 | 0.410 | 0.438 | 0.385 | 0.054 |
|  |  | 2 | 11 | 0.410 | 0.342 | 0.355 | 0.013 |
|  |  | 3 | 2 | 0.366 | 0.473 | 0.417 | 0.056 |
|  | *LSTM+ AdaSyn* | 1 | 18 | 0.337 | 0.318 | 0.411 | 0.093 |
|  |  | 2 | 6 | 0.413 | 0.395 | 0.353 | 0.042 |
|  |  | 3 | 33 | 0.408 | 0.305 | 0.343 | 0.038 |
|  | *LSTM+ Reweight* | 1 | 63 | 0.568 | 0.436 | 0.429 | 0.007 |
|  |  | 2 | 58 | 0.559 | 0.428 | 0.431 | 0.003 |
|  |  | 3 | 66 | 0.561 | 0.429 | 0.424 | 0.005 |
| eICU | *LSTM* | 1 | 81 | 0.476 | 0.277 | 0.280 | 0.003 |
|  |  | 2 | 87 | 0.510 | 0.274 | 0.283 | 0.009 |
|  |  | 3 | 86 | 0.492 | 0.279 | 0.276 | 0.003 |
|  | *LSTM+ SMOTE* | 1 | 49 | 0.385 | 0.495 | 0.410 | 0.085 |
|  |  | 2 | 55 | 0.402 | 0.445 | 0.391 | 0.055 |
|  |  | 3 | 42 | 0.433 | 0.412 | 0.471 | 0.059 |
|  | *LSTM+ AdaSyn* | 1 | 39 | 0.320 | 0.410 | 0.419 | 0.009 |
|  |  | 2 | 38 | 0.376 | 0.454 | 0.499 | 0.045 |
|  |  | 3 | 35 | 0.336 | 0.439 | 0.468 | 0.029 |
|  | *LSTM+ Reweight* | 1 | 99 | 0.473 | 0.532 | 0.526 | 0.006 |
|  |  | 2 | 90 | 0.495 | 0.512 | 0.539 | 0.026 |
|  |  | 3 | 89 | 0.454 | 0.494 | 0.502 | 0.008 |

**Supplementary Table 3:**
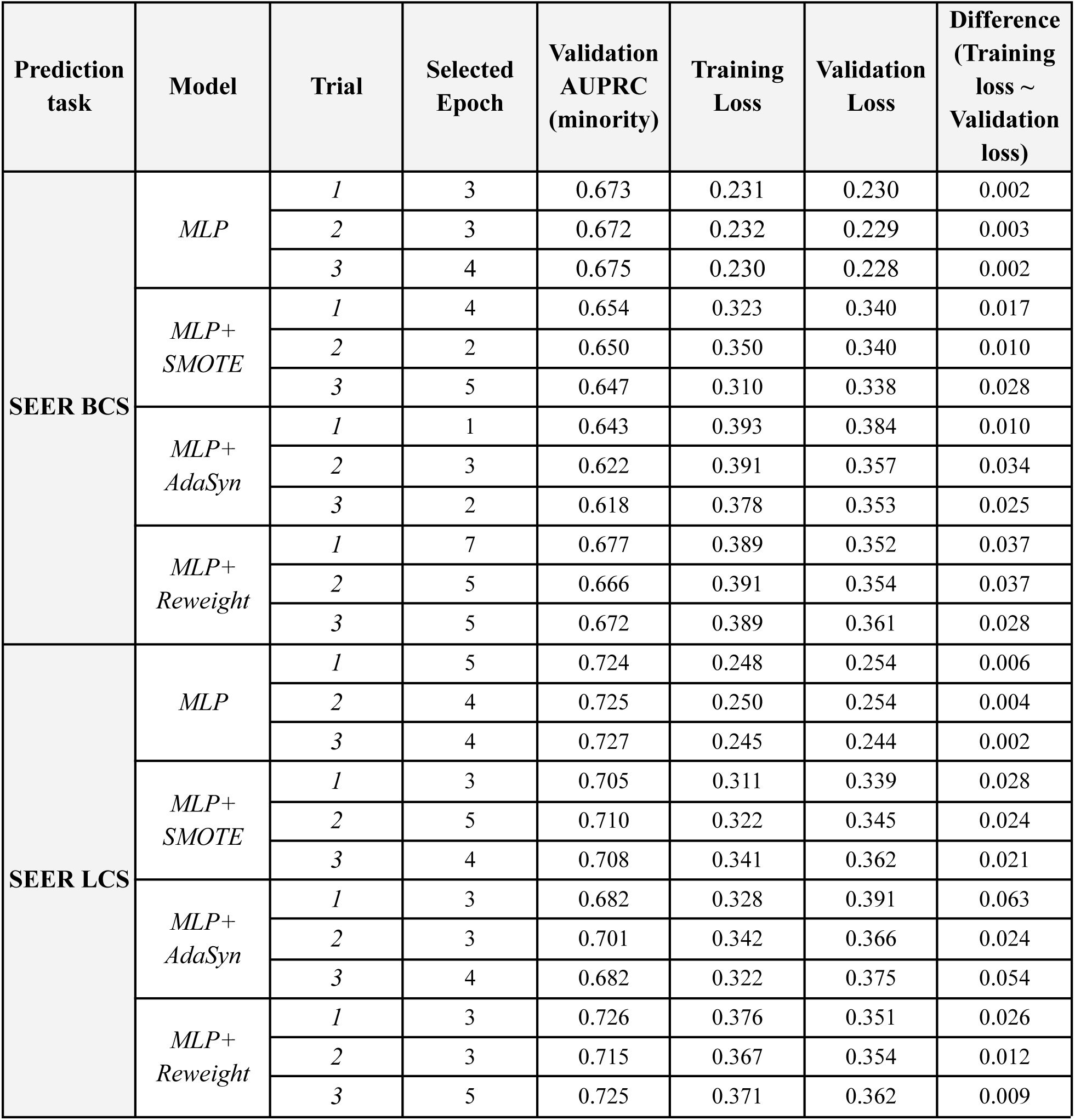
Training and validation losses of selected cancer survivability predictor models.

**Supplementary Table 4:**
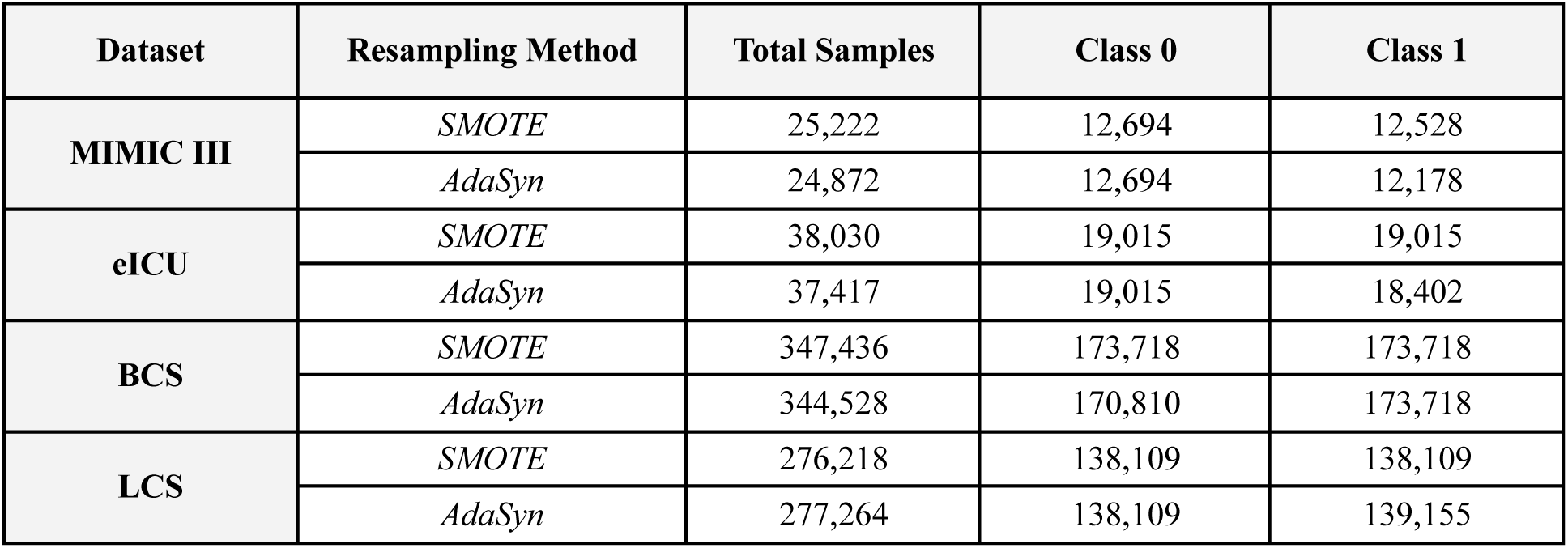
Number of samples in the training set after resampling.

| Dataset | Resampling Method | Total Samples | Class 0 | Class 1 |
| --- | --- | --- | --- | --- |
| MIMIC III | *SMOTE* | 25,222 | 12,694 | 12,528 |
|  | *AdaSyn* | 24,872 | 12,694 | 12,178 |
| eICU | *SMOTE* | 38,030 | 19,015 | 19,015 |
|  | *AdaSyn* | 37,417 | 19,015 | 18,402 |
| BCS | *SMOTE* | 347,436 | 173,718 | 173,718 |
|  | *AdaSyn* | 344,528 | 170,810 | 173,718 |
| LCS | *SMOTE* | 276,218 | 138,109 | 138,109 |
|  | *AdaSyn* | 277,264 | 138,109 | 139,155 |

**Supplementary Table 5:** Cost-sensitive learning balanced class weights.

| Dataset | Class 0 | Class 1 |
| --- | --- | --- |
| MIMIC III | 0.578 | 3.694 |
| eICU LSTM | 0.565 | 4.363 |
| LCS MLP | 0.595 | 3.122 |
| BCS MLP | 3.936 | 0.573 |

**Supplementary Table 6:** Thresholds of various models. Thresholds are identified through the validation process.

| Models/<br>Datasets | NN | NN+<br>SMOTE | NN +<br>AdaSyn | NN+<br>Reweight | XGBoost | Random<br>Forest | Ada-<br>Boost | KNN | Naive<br>Bayes |
| --- | --- | --- | --- | --- | --- | --- | --- | --- | --- |
| **eICU**<br>(NN: LSTM) | 0.24 | 0.11 | 0.14 | 0.24 | 0.22 | 0.24 | 0.24 | 0.21 | 0.17 |
| **MIMIC-III**<br>(NN: LSTM) | 0.22 | 0.19 | 0.18 | 0.22 | 0.26 | 0.25 | 0.18 | 0.21 | 0.06 |
| **SEER-BCS**<br>(NN: MLP) | 0.71 | 0.694 | 0.695 | 0.69 | 0.67 | 0.70 | 0.70 | 0.71 | 0.83 |
| **SEER-LCS**<br>(NN: MLP) | 0.34 | 0.29 | 0.31 | 0.31 | 0.32 | 0.34 | 0.32 | 0.31 | 0.11 |

**Supplementary Table 7:** Mortality risk prediction of two medical experts on attribute-varying test cases. Doctors were given below and told the attribute values may fluctuate.

| Case | Attribute values | Predicted risk (0-1) of attribute-varying test cases |  |  |  |
| --- | --- | --- | --- | --- | --- |
|  |  | Risk score by doctor 1 | Risk score by doctor 2 | Doctors' avg | Risk by MIMIC III LSTM model |
| 1 | Systolic blood pressure (mm Hg): 52<br>Diastolic blood pressure (mm Hg): 48<br>Glucose level (mg/dL): 22<br>Respiratory rate (BPM): 37<br>Heart rate (BPM): 21<br>Oxygen saturation (%): 34 | 0.75 | 0.95 | 0.85 | 0.084 |
| 2 | Systolic blood pressure (mm Hg): 79<br>Diastolic blood pressure (mm Hg): 35<br>Glucose level (mg/dL): 47<br>Respiratory rate (BPM): 41<br>Heart rate (BPM): 20<br>Oxygen saturation (%): 43 | 0.80 | 0.90 | 0.85 | 0.137 |
| 3 | Systolic blood pressure (mm Hg): 61<br>Diastolic blood pressure (mm Hg): 32<br>Glucose level (mg/dL): 35<br>Respiratory rate (BPM): 42<br>Heart rate (BPM): 48<br>Oxygen saturation (%): 26 | 0.85 | 0.90 | 0.875 | 0.611 |
| 4 | Systolic blood pressure (mm Hg): 61<br>Diastolic blood pressure (mm Hg): 43<br>Glucose level (mg/dL): 35<br>Respiratory rate (BPM): 42<br>Heart rate (BPM): 48<br>Oxygen saturation (%): 20 | 0.85 | 0.90 | 0.875 | 0.56 |
| 5 | Systolic blood pressure (mm Hg): 223<br>Diastolic blood pressure (mm Hg): 153<br>Glucose level (mg/dL): 253<br>Respiratory rate (BPM): 30<br>Heart rate (BPM): 117<br>Oxygen saturation (%): 68 | 0.50 | 0.50 | 0.50 | 0.117 |
| 6 | Systolic blood pressure (mm Hg): 295<br>Diastolic blood pressure (mm Hg): 153<br>Glucose level (mg/dL): 253<br>Respiratory rate (BPM): 45<br>Heart rate (BPM): 136<br>Oxygen saturation (%): 62 | 0.55 | 0.60 | 0.575 | 0.455 |

**Supplementary Table 8:** Deteriorating test cases labeling of mortality risk by 2 medical doctors.

| Attribute name | Case | Predicted risk (0-1) of deteriorating test cases |  |  |  |
| --- | --- | --- | --- | --- | --- |
|  |  | Medical Doctor 1 | Medical Doctor 2 | Doctors' avg | MIMIC III LSTM model (avg) |
| Respiratory rate | 1 | 0.85 | 0.60 | 0.73 | 0.13 |
|  | 2 | 0.90 | 0.80 | 0.85 | 0.19 |
|  | 3 | 0.90 | 0.60 | 0.75 | 0.16 |
| Temperature | 1 | 0.90 | 0.40 | 0.65 | 0.18 |
|  | 2 | 0.50 | 0.35 | 0.43 | 0.17 |
|  | 3 | 0.40 | 0.30 | 0.35 | 0.18 |
| Systolic BP | 1 | 0.95 | 0.90 | 0.93 | 0.13 |
|  | 2 | 0.95 | 0.90 | 0.93 | 0.03 |
|  | 3 | 0.95 | 0.90 | 0.93 | 0.20 |
| Multi-attribute | 1 | 0.80 | 0.90 | 0.85 | 0.20 |
|  | 2 | 0.85 | 0.90 | 0.88 | 0.18 |
|  | 3 | 0.80 | 0.90 | 0.85 | 0.19 |

**Supplementary Table 9:** The table shows the gradient-based test case generation steps and prediction. Each step of gradient ascent creates a new test case by changing a single attribute value. Mortality risk (MR) is predicted by three models which are trained on the same MIMIC-III training dataset.

| Attribute | Deteriorating test case | Step factor | # of Steps | Seed attribute value | Final attribute value | MR by LSTM 1 | MR by LSTM 2 | MR by LSTM 3 | MR by LR | MR by CW-LS TM |
| --- | --- | --- | --- | --- | --- | --- | --- | --- | --- | --- |
| Diastolic BP (mm Hg) | 1 | 0.01 | 16 | 52.94 | 10.77 | 0.04 | 0.22 | 0.22 | 0.03 | 0.05 |
|  | 2 | 0.01 | 100 | 55.89 | 1.19 | 0.10 | 0.04 | 0.08 | 0.20 | 0.02 |
|  | 3 | 0.01 | 100 | 89.37 | 4.49 | 0.07 | 0.06 | 0.09 | 0.14 | 0.05 |
| Systolic BP (mm Hg) | 1 | 0.2 | 54 | 119.85 | 95.00 | 0.14 | 0.22 | 0.06 | 0.02 | 0.27 |
|  | 2 | 0.2 | 35 | 124.70 | 105.35 | 0.07 | 0.11 | 0.22 | 0.13 | 0.19 |
|  | 3 | 0.2 | 170 | 125.59 | 67.99 | 0.18 | 0.21 | 0.22 | 0.05 | 0.37 |
| Resp. Rate (BPM) | 1 | 0.01 | 82 | 15.67 | 27.51 | 0.22 | 0.12 | 0.06 | 0.05 | 0.28 |
|  | 2 | 0.01 | 138 | 17.49 | 31.14 | 0.17 | 0.22 | 0.20 | 0.45 | 0.22 |
|  | 3 | 0.01 | 218 | 20.49 | 42.40 | 0.11 | 0.15 | 0.22 | 0.32 | 0.11 |
| Oxygen Sat. (%) | 1 | 0.01 | 9 | 69.26 | 0.00 | 0.03 | 0.15 | 0.15 | 0.18 | 0.01 |
|  | 2 | 0.01 | 21 | 69.83 | 0.00 | 0.03 | 0.01 | 0.03 | 0.67 | 0.01 |
|  | 3 | 0.01 | 44 | 80.68 | 0.00 | 0.02 | 0.01 | 0.01 | 0.38 | 0.03 |
| Temp (C) | 1 | 0.001 | 27 | 36.78 | 35.96 | 0.15 | 0.22 | 0.20 | 0.02 | 0.18 |
|  | 2 | 0.001 | 38 | 36.62 | 35.64 | 0.15 | 0.14 | 0.23 | 0.14 | 0.10 |
|  | 3 | 0.001 | 57 | 36.79 | 35.41 | 0.15 | 0.17 | 0.23 | 0.05 | 0.13 |

**Supplementary Table 10:** Number of test cases (excluding deteriorating condition) in one test set created from a single seed case in the original MIMIC-III dataset. Five test sets are produced for each attribute or combination of attributes using five different seeds. The deteriorating conditions test is generated by three seeds resulting in 12 test cases.

| Test type | Tested attribute | Number of cases per set | Number of total cases |
| --- | --- | --- | --- |
| GCS test | *GCS total* | 120 | 600 |
|  | *GCS eye-motor* | 24 | 120 |
|  | *GCS eye-verbal* | 20 | 100 |
|  | *GCS motor-verbal* | 30 | 150 |
| GCS test total case |  | 194 | 970 |
| Single attribute critical zone test | *Diastolic BP* | 250 | 1,250 |
|  | *Glucose* | 600 | 3,000 |
|  | *Oxygen saturation* | 101 | 505 |
|  | *Respiratory rate* | 101 | 505 |
|  | *Systolic BP* | 350 | 1,750 |
|  | *Temperature* | 13 | 65 |
| Critical zone test total case |  | 1415 | 7,075 |
| Double-attribute critical zone test | *Diastolic BP & Glucose* | 2,500 | 12,500 |
|  | *Diastolic BP & Systolic BP* | 3,500 | 17,500 |
|  | *Respiratory rate & Heart rate* | 2,500 | 12,500 |
| Double attribute critical zone test total case |  | 8,500 | 42,500 |
| Multi-attribute critical zone test (6 attributes) | *High zone* | 12,695 | 63,475 |
|  | *Low zone* | 12,695 | 63,475 |
| Multi-attribute critical zone test (6 attributes) total case |  | 25,390 | 126,950 |
| Deteriorating conditions test (gradient ascent method) | *Single attribute* | 9 | 9 |
|  | *Triple-attribute* | 3 | 3 |
| Deteriorating conditions test total case |  | 12 | 12 |
| Grand total |  | 35,511 | 177,507 |

**Supplementary Table 11:**
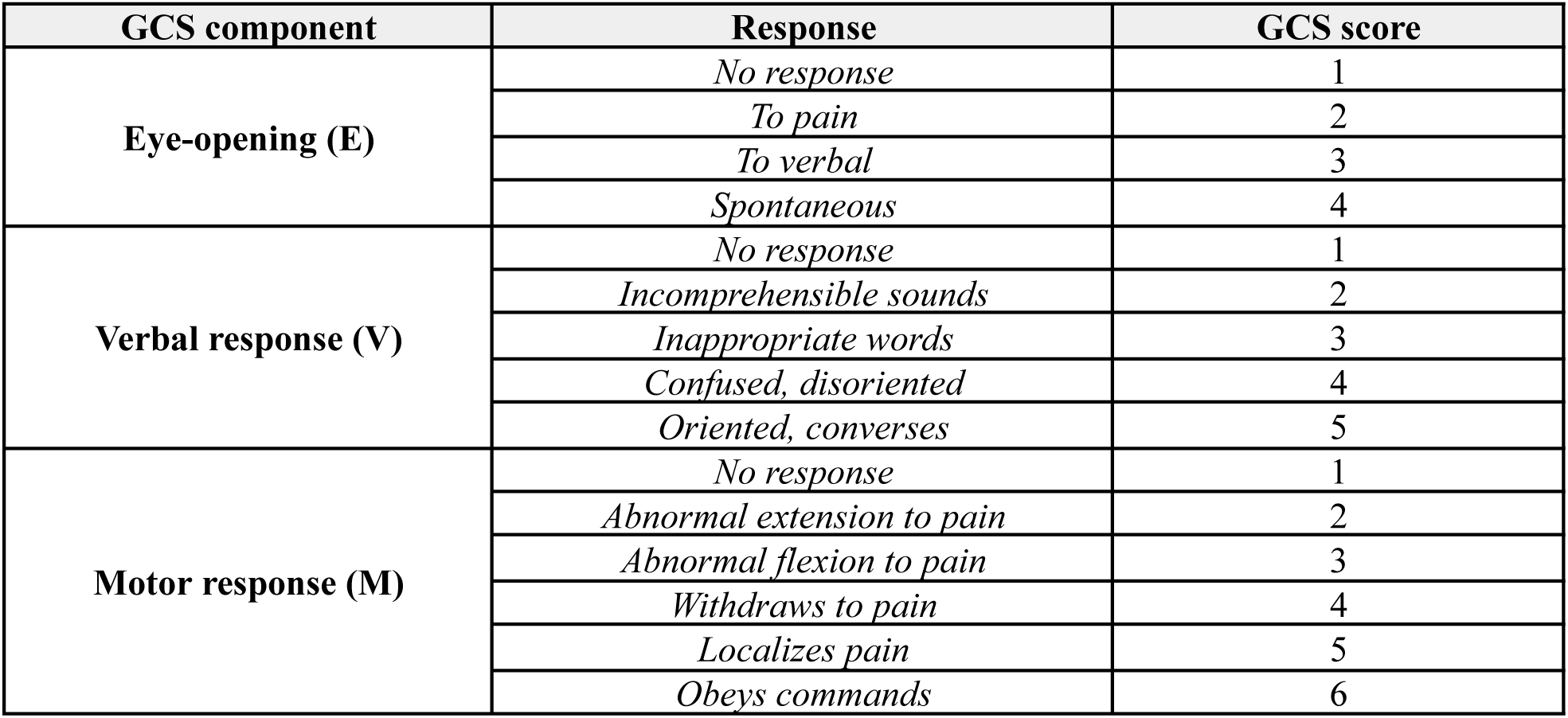
Clinical Assessment of Neurological Function: Glasgow Coma Scale (GCS) Scoring System.

| **GCS component** | **Response** | **GCS score** |
| --- | --- | --- |
| **Eye-opening (E)** | *No response* | 1 |
|  | *To pain* | 2 |
|  | *To verbal* | 3 |
|  | *Spontaneous* | 4 |
| **Verbal response (V)** | *No response* | 1 |
|  | *Incomprehensible sounds* | 2 |
|  | *Inappropriate words* | 3 |
|  | *Confused, disoriented* | 4 |
|  | *Oriented, converses* | 5 |
| **Motor response (M)** | *No response* | 1 |
|  | *Abnormal extension to pain* | 2 |
|  | *Abnormal flexion to pain* | 3 |
|  | *Withdraws to pain* | 4 |
|  | *Localizes pain* | 5 |
|  | *Obeys commands* | 6 |

**Supplementary Table 12:**
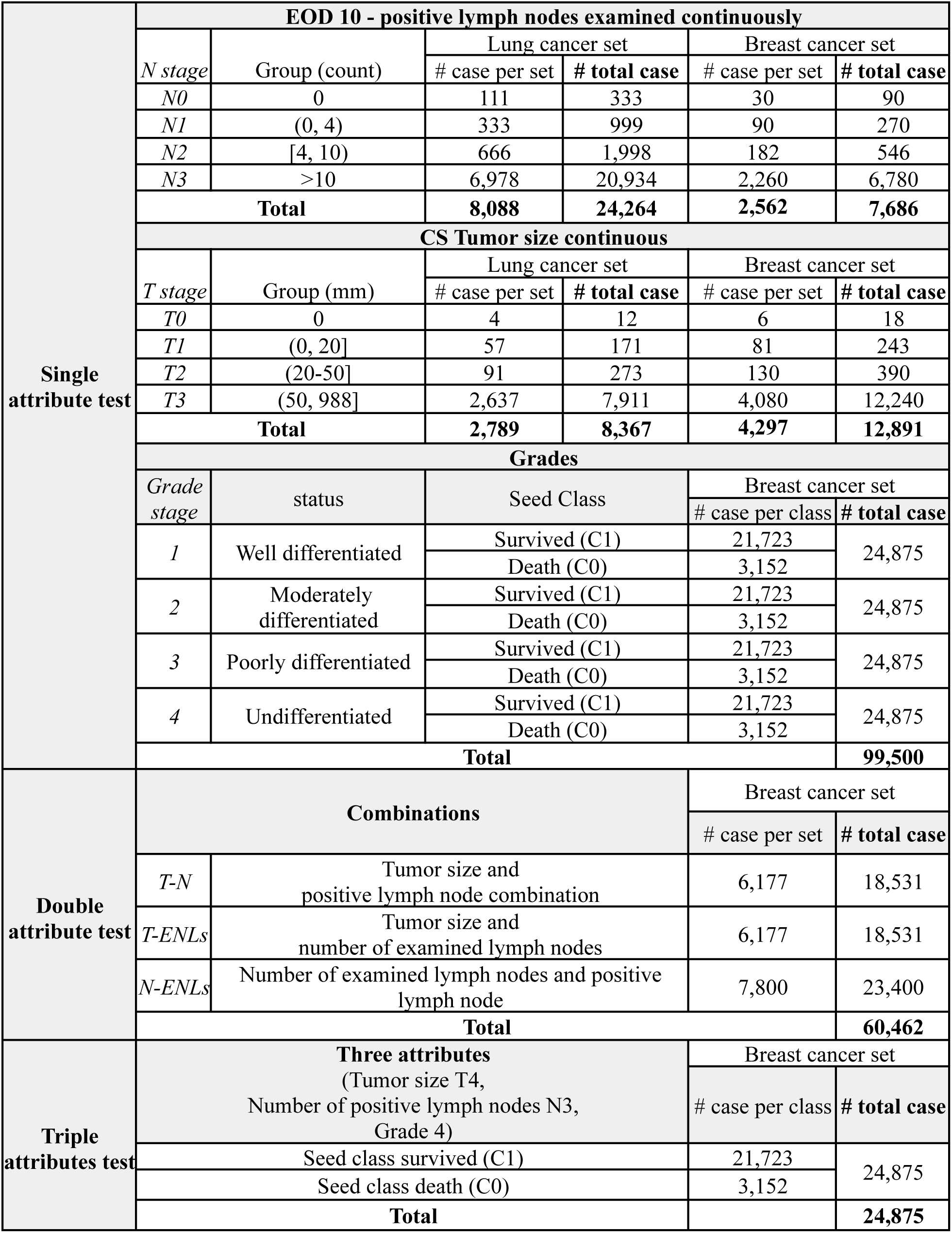
Data distribution of created test set for SEER 5-year breast cancer survivability (BCS) and lung cancer survivability (LCS).

**Supplementary Table 13:**
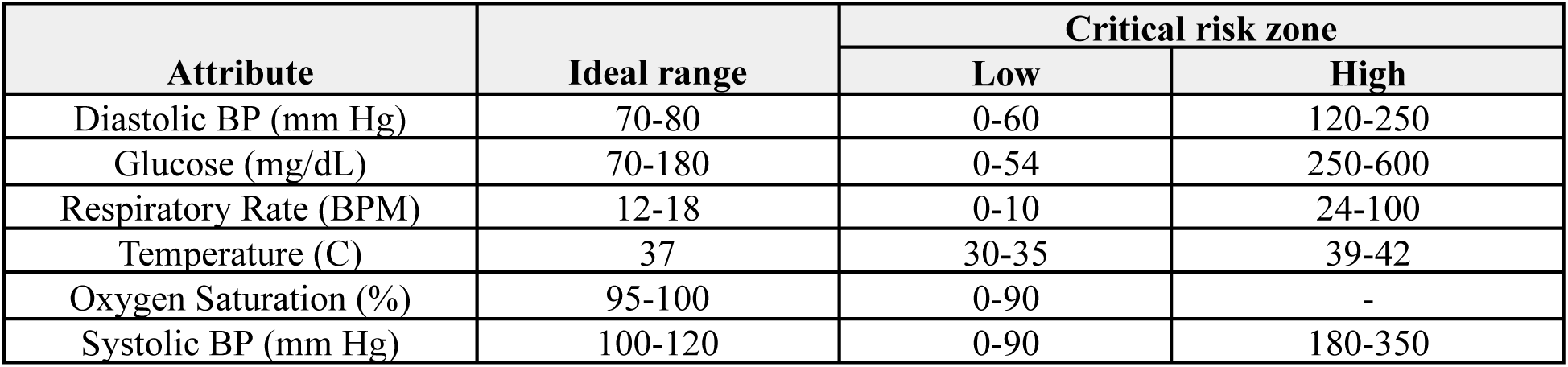
Ranges of important vitals.

**Supplementary Table 14:** Mean standard deviation of the attribute of the original MIMIC-III dataset.

|  | All |  | Class 0 (Non-death) |  | Class 1 (Death) |  |
| --- | --- | --- | --- | --- | --- | --- |
|  | Mean | SD (avg) | Mean | SD (avg) | Mean | SD (avg) |
| **Diastolic bp (mm Hg)** | 61.45 | 15.61 | 61.60 | 14.20 | 60.46 | 24.63 |
| **GCS eye-opening** | 3.22 | 0.59 | 3.34 | 0.58 | 2.47 | 0.63 |
| **GCS motor response** | 5.33 | 0.61 | 5.46 | 0.60 | 4.47 | 0.70 |
| **GCS total** | 11.85 | 1.65 | 12.28 | 1.67 | 9.24 | 1.51 |
| **GCS verbal response** | 3.32 | 0.67 | 3.48 | 0.71 | 2.30 | 0.46 |
| **Glucose (mg/dL)** | 141.54 | 48.34 | 140.72 | 49.67 | 146.81 | 39.78 |
| **Heart Rate (BPM)** | 86.67 | 10.08 | 86.14 | 9.91 | 90.03 | 11.13 |
| **Oxygen saturation (%)** | 97.98 | 11.50 | 98.23 | 12.61 | 96.44 | 4.39 |
| **Respiratory rate (BPM)** | 20.96 | 19.92 | 21.05 | 22.27 | 20.44 | 4.95 |
| **Systolic bp (mm Hg)** | 120.25 | 15.67 | 120.78 | 15.48 | 116.85 | 16.92 |
| **Temperature (C)** | 36.94 | 0.71 | 36.95 | 0.69 | 36.88 | 0.86 |

**Supplementary Table 15:** Selected seeds from the MIMIC-III dataset. These five seeds are used to create attribute-based test cases. Mean and SD represent the average and standard deviation values of the particular attribute. Missing values were ignored for calculating the mean and standard deviation.

| Seeds | 1 |  | 2 |  | 3 |  | 4 |  | 5 |  |
| --- | --- | --- | --- | --- | --- | --- | --- | --- | --- | --- |
| Attribute | *Mean* | *SD* | *Mean* | *SD* | *Mean* | *SD* | *Mean* | *SD* | *Mean* | *SD* |
| **Diastolic BP (mm Hg)** | 55.40 | 8.27 | 55.85 | 7.00 | 55.77 | 4.29 | 66.08 | 5.39 | 66.72 | 10.05 |
| **GCS eye opening** | 4.00 | 0.00 | 4.00 | 0.00 | 4.00 | 0.00 | 4.00 | 0.00 | 4.00 | 0.00 |
| **GCS motor response** | 6.00 | 0.00 | 6.00 | 0.00 | 6.00 | 0.00 | 6.00 | 0.00 | 6.00 | 0.00 |
| **GCS total** | 15.00 | 0.00 | 15.00 | 0.00 | 15.00 | 0.00 | 15.00 | 0.00 | 15.00 | 0.00 |
| **GCS verbal response** | 5.00 | 0.00 | 5.00 | 0.00 | 5.00 | 0.00 | 5.00 | 0.00 | 5.00 | 0.00 |
| **Glucose (mg/dL)** | 118.75 | 13.84 | 153.70 | 22.60 | 97.00 | 8.19 | 126.50 | 7.33 | 109.00 | 21.21 |
| **Heart Rate (BPM)** | 74.27 | 8.29 | 64.40 | 5.72 | 66.90 | 6.22 | 99.34 | 9.89 | 67.67 | 5.84 |
| **Mean BP (mm Hg)** | 73.15 | 5.69 | 84.67 | 7.32 | 72.64 | 4.63 | 84.88 | 4.87 | 84.92 | 10.53 |
| **Oxygen sat. (%)** | 96.71 | 1.68 | 96.81 | 1.13 | 98.51 | 2.01 | 99.06 | 0.91 | 95.10 | 1.79 |
| **Respiratory rate (BPM)** | 15.33 | 3.87 | 16.33 | 3.50 | 17.07 | 2.11 | 15.52 | 3.38 | 14.22 | 2.83 |
| **Systolic BP (mm Hg)** | 108.00 | 8.91 | 116.34 | 12.62 | 106.38 | 6.54 | 122.48 | 5.96 | 121.32 | 14.61 |
| **Temperature (C)** | 37.38 | 0.37 | 36.97 | 0.56 | 37.41 | 0.27 | 37.21 | 0.27 | 37.52 | 0.50 |

**Supplementary Table 16:** Average accuracy of machine learning models under various testing conditions for in-hospital mortality prediction.

|  |  | Accuracy |  |  |
| --- | --- | --- | --- | --- |
|  | Attributes | LSTM | CW-LSTM | LR |
| **Single attribute critical zone tests** | *Respiratory rate* | 64.67% | 13.00% | 59.00% |
|  | *Temperature* | 58.97% | 23.08% | 46.15% |
|  | *Diastolic BP* | 23.60% | 23.60% | 23.60% |
|  | *Oxygen saturation* | 10.89% | 10.89% | 38.61% |
|  | *Systolic BP* | 35.24% | 30.00% | 25.43% |
|  | *Glucose* | 33.62% | 33.62% | 33.62% |
|  | **Average accuracy of single-attribute tests** | **37.83%** | **22.36%** | **37.74%** |
| **GCS** |  | 33.33% | 33.33% | 100% |
| **Multiple attribute critical zone tests** | **Zones** |  |  |  |
|  | *High critical zone* | 22.50% | 40.10% | 0.15% |
|  | *Low critical zone* | 68.80% | 98.40% | 12.30% |
| **Average accuracy of multi-attribute tests** |  | **45.65%** | **69.25%** | **6.23%** |
| **Deteriorating condition tests** | Any critical zones | 83.33% | 66.66% | 33.33% |

**Supplementary Table 17:**
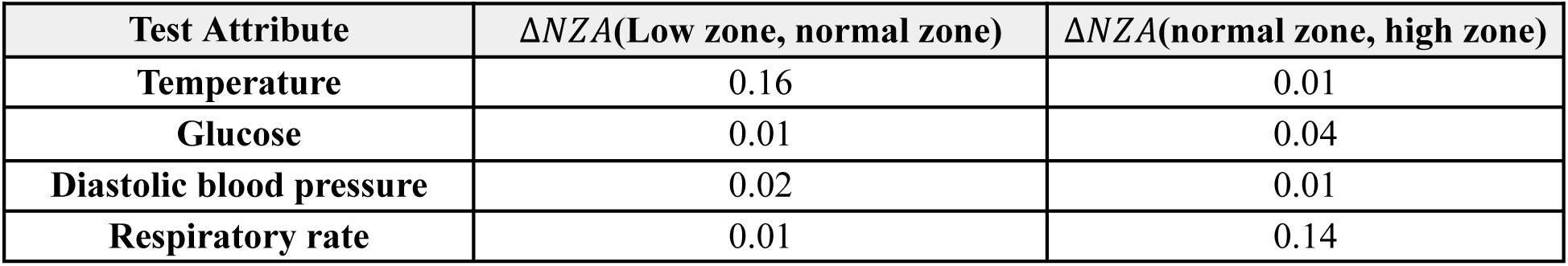
Changes in neural zone activation values of the LSTM model. Values in columns 2 and 3 represent the average difference of a neuron’s activation score between two different regions (critically low zone, normal zone, and critically high zone).

| Test Attribute | $\Delta NZA(\text{Low zone, normal zone})$ | $\Delta NZA(\text{normal zone, high zone})$ |
| --- | --- | --- |
| Temperature | 0.16 | 0.01 |
| Glucose | 0.01 | 0.04 |
| Diastolic blood pressure | 0.02 | 0.01 |
| Respiratory rate | 0.01 | 0.14 |

**Supplementary Table 18:** Average accuracy of MLP model under different test scenarios for 5-year breast cancer survivability prediction.

|  |  |  |  |  |
| --- | --- | --- | --- | --- |
| Single attribute test | EOD 10 - positive lymph nodes examined |  |  |  |
|  | *N stage* | Group (count) | Accuracy |  |
|  | *N0* | 0 | 100% |  |
|  | *N1* | (0, 4) | 0% |  |
|  | *N2* | [4, 10) | 0% |  |
|  | *N3* | >10 | 74.4% |  |
|  | Average |  | 43.6% |  |
|  | CS Tumor size |  |  |  |
|  | *T stage* | Group (mm) | Accuracy |  |
|  | *T0* | 0 | 100% |  |
|  | *T1* | (0, 20] | 0% |  |
|  | *T2* | (20-50] | 0% |  |
|  | *T3* | (50, 988] | 0% |  |
|  | Average |  | 25% |  |
|  | Grades |  |  |  |
|  | *Grade stage* | status | Seed Class | Accuracy |
|  | 1 | Well-differentiated | Survived (C1) | 96.97% |
|  |  |  | Death (C0) | 48.67% |
|  | 2 | Moderately differentiated | Survived (C1) | 4.55% |
|  |  |  | Death (C0) | 57.93% |
|  | 3 | Poorly differentiated | Survived (C1) | 7.44% |
|  |  |  | Death (C0) | 65.77% |
|  | 4 | Undifferentiated | Survived (C1) | 6.66% |
|  |  |  | Death (C0) | 63.86% |
|  | Average |  | 43.98% |  |
| Double attribute test | Double Combinations |  | Accuracy |  |
|  | T-N: Tumor size and positive lymph node combination |  | 92.97% |  |
|  | T-ENL: Tumor size and number of examined lymph nodes |  | 0% |  |
|  | N-ENL: Number of examined lymph nodes and positive lymph node |  | 19.6% |  |
|  | Average |  | 37.52% |  |
| Triple attributes test | Three attributes<br>(Tumor size T4, Number of positive lymph nodes N3, Grade 4) |  | Accuracy |  |
|  | Seed class survived (C1) |  | 89.89% |  |
|  | Seed class death (C0) |  | 98.92% |  |
|  | Average |  | 94.41% |  |

**Supplementary Table 19:** Wasserstein distances between various training and testing datasets.

| **Training data** | **Testing data** | **Wasserstein distances (average)** |
| --- | --- | --- |
| *Original MIMIC-III train set* | *Original MIMIC-III test set* | 12.42 |
| *Original MIMIC-III train set* | *Synthesized MIMIC-III multi-attribute test set* | 33.38 |
| *Original SEER BCS train set* | *Original SEER BCS test set* | 2.1 |
| *Original SEER BCS train set* | *Synthesized SEER BCS triple-attribute test set* | 9.75 |

